# Harnessing confounding and genetic pleiotropy to identify causes of disease through proteomics and Mendelian randomisation – ‘MR Fish’

**DOI:** 10.1101/2024.07.11.24310200

**Authors:** Alasdair N Warwick, Aroon D Hingorani, Anthony P Khawaja, María Gordillo-Marañón, Abraham Olvera-Barrios, Kelsey V Stuart, Catherine Egan, Adnan Tufail, Reecha Sofat, Valerie Kuan Po Ai, Chris Finan, Amand F Schmidt

**Affiliations:** UCL Institute of Cardiovascular Science, University College London, London, UK; NIHR Biomedical Research Centre, Moorfields Eye Hospital NHS Foundation Trust & University College London Institute of Ophthalmology, London, UK; UCL British Heart Foundation Research Accelerator; London, UK; Department of Pharmacology and Therapeutics, University of Liverpool, Liverpool, UK; British Heart Foundation Data Science Centre, Health Data Research UK, London, UK; Department of Cardiology, Amsterdam Cardiovascular Sciences, Amsterdam University Medical Centres, University of Amsterdam; Amsterdam, the Netherlands

## Abstract

We propose an extension of the Mendelian randomisation (MR) paradigm (‘MR-Fish’) in which the confounded disease association of an index protein (‘the bait’) is harnessed to identify the causal role of different proteins (‘the catch’) for the same disease. Using C-reactive protein (CRP) as the bait, *cis*-MR analyses refuted a causal relationship of CRP with a wide range of diseases that associate with CRP in observational studies, including type 2 diabetes (T2DM) and coronary heart disease (CHD), suggesting these associations are confounded. Using ‘MR-Fish’, and leveraging large-scale proteomics data, we find evidence of a causal relationship with multiple diseases for several proteins encoded by genes that are *trans* hits in genome wide association analysis of CRP. These include causal associations of IL6R and FTO with CHD and T2DM; as well as ZDHHC18 with several circulating blood lipid fractions. Among the proteins encoded by genes that are *trans*-for-CRP we identified 28 that are druggable. Our findings point to a general approach using MR analysis with proteomics data to identify causal pathways and therapeutic targets from non-causal observational associations of an index protein with a disease.

## Introduction

Observational studies have identified associations between many circulating biomarkers and diseases.^1^ Proteins represent an important subcategory of circulating biomarkers because they are the effector molecules in biology and the targets of most drugs. For example, in a recent umbrella review, a higher circulating concentration of the inflammatory biomarker C-reactive protein (CRP) was consistently found to associate with a higher risk of coronary heart disease (CHD), type 2 diabetes, certain cancers, dementia and several other adverse health outcomes.^2^

However, observational associations between a biomarker and disease may not be causal but arise due to a confounder: a measured, unmeasured, or unknown factor that associates causally with both the biomarker and disease of interest (**Fig. 1**). For example, in the association of CRP with CHD, potential confounders include smoking, blood pressure, obesity, diabetes, and other agents of the inflammatory response such as interleukin-6. All are associated with CRP, and some, e.g. blood pressure and diabetes, are proven causes of CHD.^2^

**Fig. 1.**
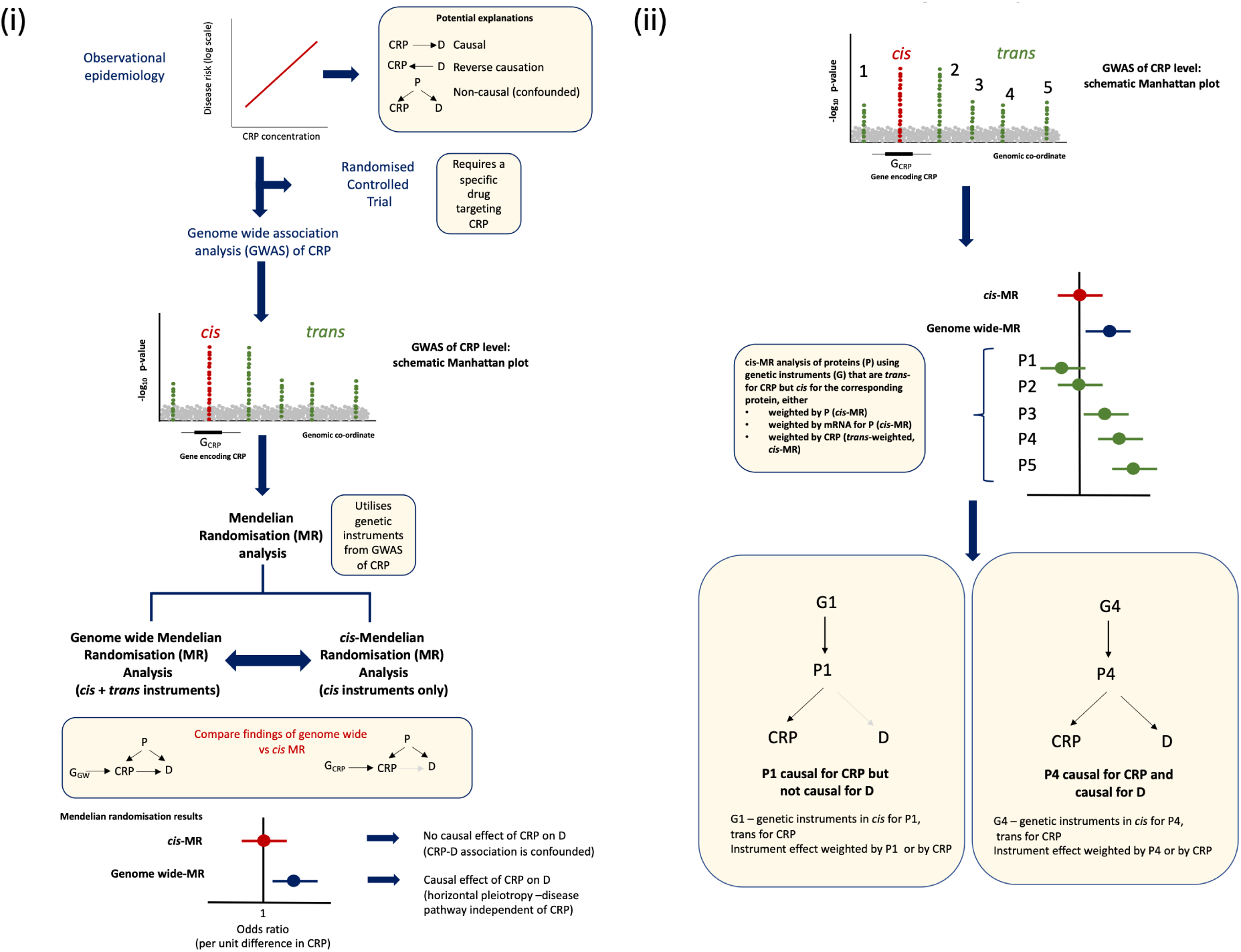
Overview of the conceptual basis of MR-Fish, using C-reactive protein (CRP) as an example of a ‘bait’ protein. (i) Observational associations between CRP and disease (D) may be causal or arise due to confounding. Confounding would occur in the presence of other (often unidentified) proteins that are causally associated with both CRP and D. Whether the CRP-D association is causal could be confirmed or refuted in a randomised controlled trial of a CRP-lowering or blocking drug (if available), or by performing a Mendelian randomisation (MR) analysis. Genetic instruments for an MR analysis of CRP may be selected from throughout the genome (genome-wide MR, including variants that are *trans*-for-CRP) or be restricted to instruments at the locus encoding CRP (*cis*-MR). In genome-wide MR, the inclusion of variants outside the encoding gene (*trans*-for-CRP) may introduce horizontal pleiotropy. This may result in a discrepancy in causal inferences between *cis*-MR, which is relatively protected from horizontal pleiotropy, and genome-wide MR. (ii) MR-Fish seeks to identify proteins that are potential confounders of the CRP-D association. A GWAS of CRP may provide a source of potential confounders because the loci identified that are *trans* for CRP encode proteins that are causally associated with CRP (one of the criteria for being a confounder). The causal relevance of these *trans*-for-CRP proteins for D may then be assessed using *cis*-MR weighted either by the encoded protein P or mRNA for P (utilising proteomic or transcriptomic GWAS respectively), or alternatively *trans*-weighted by CRP (i.e. drawing on the original GWAS for CRP levels), In the example shown, P4 is a confounder of the CRP-D association and, by definition, is therefore causal for D, whereas P1 is not. Abbreviations: CRP = C-reactive protein; D = Disease; G = Gene; GWAS = Genome-wide association study; MR = Mendelian randomisation; mRNA = messenger ribonucleic acid; OR = Odds ratio; P = Protein.

Statistical adjustment can be made for measured confounders in observational studies but residual confounding remains a problem because not all confounders are known, and many are measured imperfectly meaning they cannot be fully adjusted for.^3,4^ A randomised controlled trial of a selective drug intervention, in the current example to reduce the concentration or block the effects of CRP,^5^ would distinguish causal from confounded associations of CRP with a given disease. However, drug development for a single protein target is costly^6,7^ and extending this approach to the many thousands of protein-disease associations is impractical.

A scalable alternative to prioritising therapeutic targets is to use genetic variants that influence the expression or action of a protein to estimate its causal effect on a disease.^8^ The genetic effect estimate should be unbiased (provided certain assumptions are met) because, analogous to treatment allocation in a clinical trial, genetic variation is determined by a randomised allocation at conception (Mendelian randomisation; MR) which reduces confounding.^9–12^

A genome-wide association study (GWAS) of protein levels can identify genetic variants associated with protein expression, and these variants can be used as instruments in MR analysis to model the effect of targeting the protein with a drug. The instruments employed could be selected from within or around the encoding gene (said to be acting in *cis*) or from throughout the genome (genome-wide), including variants located outside the encoding gene (acting in *trans*) (**Fig. 1, Mathematical appendix Table A1**).^13,14^ We have argued previously that variants acting in *cis* are more likely to fulfil critical assumptions of the MR approach than are variants acting in *trans*, making *cis*-MR the preferred analytical approach where a protein is the biomarker of interest (see **Mathematical appendix Table A1**).^13,15^ The reason, based on Crick’s Central Dogma, is that the effect of a *cis* acting genetic variant is most likely to be mediated through rather than independently of the encoded protein of interest, upholding the so-called exclusion restriction assumption. This assumption states that the genetic instrument used in an MR analysis should be associated with disease only through the exposure of interest.^16^ In contrast, a genetic variant that is *trans* to the protein of interest has the potential to affect disease risk directly, through the protein encoded by the gene at that locus or via a pathway that may bypass the protein of interest, a phenomenon referred to as horizontal pleiotropy (**Fig. 1, Mathematical appendix Table A1**).^13,15^

Confounding in observational epidemiology and horizontal pleiotropy are two views on the same phenomenon. The former compromises causal inference in observational studies and the latter in MR analysis. To illustrate the point using horizontal pleiotropy as an example, variants in the *IL6R* gene encoding the interleukin-6 receptor associate with CRP concentration (in *trans*) and with CHD,^17^ whereas variants in the CRP gene (which are *cis*-for-CRP expression), and which also associate with CRP concentration, do not associate with CHD.^18^ This suggests that CRP itself is not causal for CHD and that the interleukin-6 receptor plays a causal role in CHD through pathways that are independent of CRP. Using *IL6R* variants to infer a causal effect of CRP on CHD using MR would therefore be misleading. We refer to this type of problematic analysis, which runs a high risk of spurious causal inference as a *trans*-MR analysis of CRP on CHD to distinguish it from a *cis*-MR analysis of CRP that utilises genetic instruments from the CRP locus itself (**Fig. 1, Mathematical appendix Table A1**). The demonstration of an effect of variants in *IL6R* on both CRP and CHD suggests that interleukin-6 is one confounder of the observational association between CRP and CHD (**Fig. 1, Mathematical appendix Table A1**).

We use this logic to propose and test an extension of the MR paradigm which, for convenience, we term ‘MR-Fish’, emphasising that MR-Fish is a concept not an analytical tool. In ‘MR-Fish’, the disease association of an index protein (‘the bait’), demonstrated to be due to confounding through *cis*-MR analysis, is then harnessed to identify the causal role of different proteins (‘the catch’) for the same disease. ‘MR-Fish’ requires an observational association between the bait protein and disease (even if this is confounded); and the presence of both *cis*- and *trans*-acting genetic variants for the bait protein, which are discovered using a GWAS. We illustrate the MR-Fish approach using CRP as a bait protein. CRP provides a useful example because it has been associated with a wide range of diseases in observational studies, which suggests that either CRP is causal for these outcomes, or there are factors that confound the associations that are the true causes of disease, many of which remain to be identified. Moreover, several large GWAS of circulating CRP concentration have been conducted which have identified variants located throughout the genome that are associated with CRP concentration,^19,20^ some of which are *cis*- and some *trans*-for-CRP, providing the requisite genetic tools for the MR-Fish approach.

## Results

### Overview of the workflow

We first contrasted the findings from MR analysis of CRP using the two most employed approaches *cis*-MR and genome-wide MR. As outcomes, we examined 24 biomarkers and 22 disease outcomes reported to be associated with CRP in prior observational studies.^2^ We took *cis*-MR analysis as the gold-standard approach to assess the causal relevance of CRP for these conditions, with genome-wide MR analysis being used to scope the potential for proteins other than CRP to play a causal role for the same set of diseases and therefore to confound the association of CRP with these endpoints. Next, we used variants identified by GWAS as *trans*-for-CRP and employed them as instruments in individual *cis*-MR analyses of the proteins encoded by those genes. We weighted the effects of these *cis*-acting variants by the expression of the encoded protein itself, when the required summary effect estimates were available from a GWAS of proteins measured using the Somalogic proteomics platform, and/or by the expression of the corresponding mRNA in circulating blood cells. We refer to these analyses, where the weighting variable is the product of the encoded gene, as ‘*cis*-weighted *cis*-MR analysis’ (reduced to ‘*cis*-MR analysis’ for convenience). However, in all cases it was also possible to weight the effect of these *trans*-for-CRP genetic variants by CRP, because all loci were initially identified through a GWAS of CRP concentration. We refer to this type of analysis as ‘*trans*-weighted *cis*-MR analysis’. It is important to emphasise that in this type of analysis that although the weighting variable is a different protein (CRP; hence *trans*-weighted) the causal inference relates to the protein encoded by the gene where the genetic instruments are located, not CRP (**Fig. 2**) We provide further discussion regarding interpretation and empirical validation of the approach in the separate **Mathematical appendix**.

**Fig. 2.**
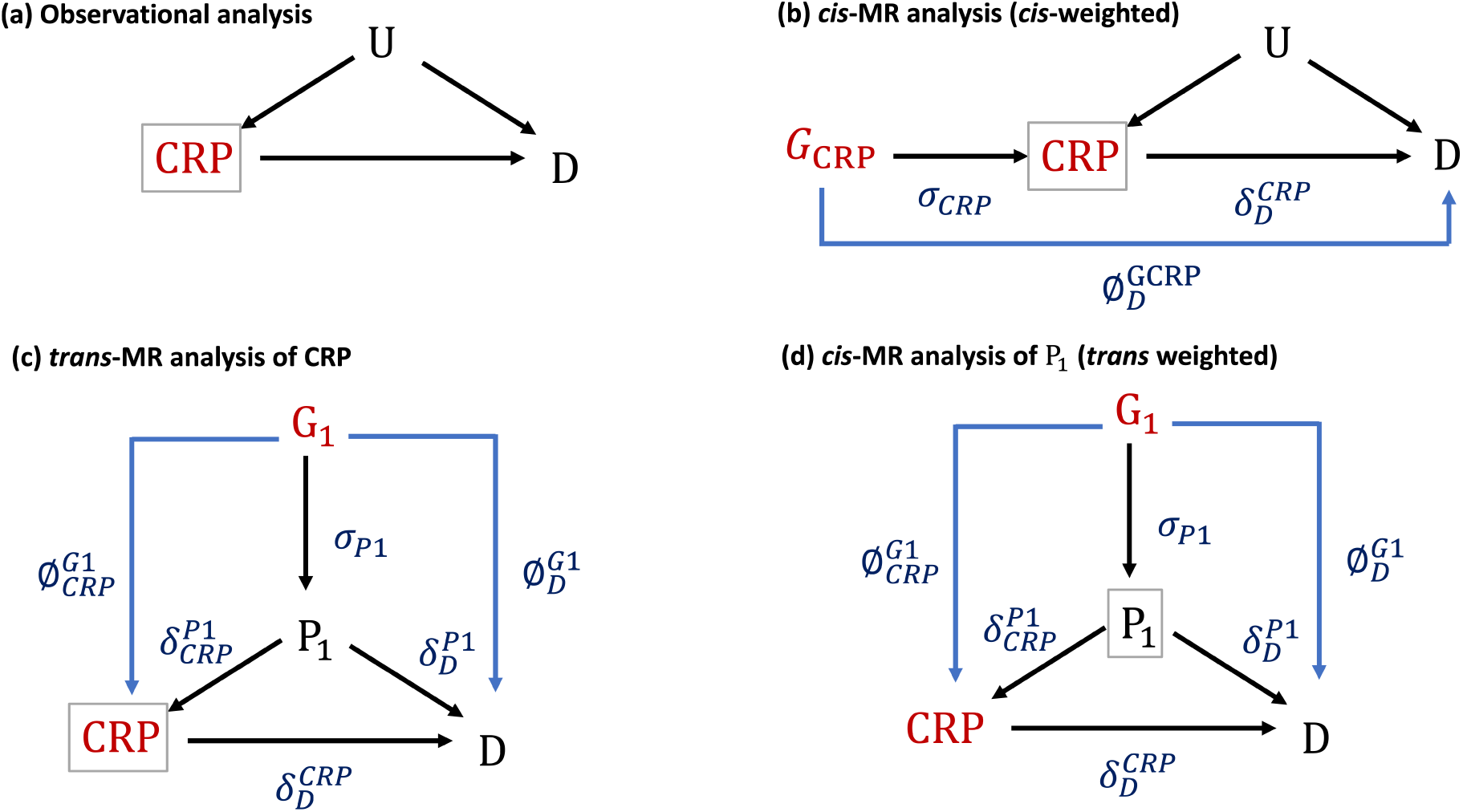
(a) Directed acyclic graph depicting an observational analysis of the relationship between the exposure (CRP) and a disease outcome *D*. The term *U* depicts confounders of the CRP - *D* association. (b and c) Directed acyclic graphs depicting alternative versions of Mendelian randomisation (MR) analysis of the exposure (CRP) and the diseases outcome *D*. Possible instruments for a MR analysis are variants in the gene *G_CRP_* encoding CRP (*cis*-MR analysis of CRP on *D*) or the gene G_1_ encoding protein P_1_, which is one confounder of the CRP - *D* association (*trans*-MR analysis of CRP on *D*). The term a refers to the effect of DNA sequence variants in a gene acting in *cis* on the encoded protein (e.g. *σ_CRP_* refers to the effect of variants in the CRP gene on CRP concentration); *δ* to the effect of a protein on an outcome (e.g. 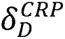 refers to the effect of CRP on disease outcome *D*); and Φ to pathways leading to horizontal pleiotropy (e.g. 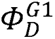 refers to horizontal pleiotropy between gene *G*_1_ and disease *D*, that might arise due to confounding by linkage disequilibrium with variants in a nearby gene). (d) Directed acyclic graph depicting a *cis*-MR analysis of the protein *P*_1_ on *D* where the genetic instruments used are *cis* for *P*_1_ but are weighted by their effect on CRP while retaining causal inference on *P*_1_ (*trans*-weighted *cis* MR analysis). In each case, a grey box surrounds the exposure of interest and the red text indicates the variable(s) measured. MR estimates under each scenario, potential sources of pleiotropy under the different MR analysis approaches, their mitigation, and other sources of bias are summarised in the **mathematical appendix, Table A1**.

### Comparison of *cis*- and genome-wide MR analysis of CRP

To evaluate the extent to which horizontal pleiotropy could bias any causal inference from genome-wide MR analysis of CRP on a variety of disease outcomes, we first compared effect estimates from *cis*-MR analysis of CRP with those from genome-wide MR analysis, which includes instruments that are *trans*-for-CRP. All analyses used CRP as the exposure of interest (based on summary statistics from the CRP GWAS of Lighart *et al.*)^19^. GWAS summary statistics for 22 disease endpoints and 24 other biomarkers associated with CRP in previous observational analyses^2,21–44^ were used as outcomes.

At a multiplicity-corrected significance threshold p<0.001 (0.05/46 outcomes), *cis*-MR analysis indicated that higher CRP was causally associated with eight biomarkers or disease endpoints (higher HDL-C concentration, monocyte count, FEV1, and FVC, as well as lower neutrophil count, and lower risk of CKD, bone fractures, and PBC). By contrast, genome-wide MR analysis of CRP using the IVW method yielded 32 significant associations at the multiplicity-corrected significance threshold p<0.001; 13 associations with disease endpoints and 19 with other biomarkers. As the aim of this analysis was to scope the potential for horizontal pleiotropy, the *cis*-MR analysis incorporated multiple genetic instruments in the vicinity of the CRP gene, selected using an algorithm that removed overinfluential variants (see Methods), whereas the genome-wide analysis of CRP utilized the ‘inverse variance weighted’ (IVW) rather than pleiotropy robust genome-wide MR methods.

We used statistical tests for interaction to identify outcomes for which the findings from *cis*-MR analysis of CRP and genome-wide MR analyses differed significantly (using a Bonferroni corrected significance threshold of p<0.05/46 tests, **Extended Data Fig. 1**). Note, the individual MRs do not have to be significant for this test. This identified 15 outcomes with significantly differing estimates. Three categories of discordance were observed (**Supplementary Table 1**): (i) for two diseases (Alzheimer’s disease and CHD) and five biomarkers (apolipoprotein (Apo)B, cognitive function, FEV1, HbA1c, and low density lipoprotein cholesterol (LDL-C)), the MR estimate from one source (*cis*-for-CRP or genome-wide) was significant (at p<0.05) but the other was not; (ii) for two diseases (CKD, PBC) and two biomarkers (FVC, HDL-C), both MR estimates were significant with the same effect direction, but differed in magnitude; and (iii) for four biomarkers (bone mineral density (BMD), FEV1/FVC ratio, leukocyte and neutrophil counts), both MR estimates were significant but with opposite effect directions.

### Identification of protein-disease relationships independent of CRP

Overall, the discordance and excess of associations from genome-wide compared to *cis*-MR analysis, suggested causal effects for genes (and their encoded proteins) that are *trans*-for-CRP (rather than CRP itself) on a range of biomarkers and disease outcomes.

To investigate this directly, we conducted individual *cis*-weighted *cis*-MR-analyses in turn for each of the 49 such proteins on the 46 evaluated outcomes that were previously associated with circulating CRP concentration through observational analysis^19^. In *cis*-MR analysis the effect of the genetic instruments selected at each locus can be weighted by their effects on circulating level of the encoded protein. However, such analyses are contingent on the requisite effect estimates being available from proteomic or transcriptomic GWAS. Alternatively, they could be weighted by the expression of the corresponding mRNA but this involves making a choice on the relevant tissue. However, in all cases, an alternative is to conduct *cis*-MR analysis weighted by CRP (i.e., *trans*-weighted *cis*-MR analysis; **Fig. 2d**). In all three approaches, the causal inference remains on the protein encoded by the gene where the instruments are located, not on CRP.

Trans-weighted cis-MR analysis of proteins encoded by genes that are *trans*-for-CRP Figure 3 illustrates the summary findings from the *trans*-weighted *cis*-MR analysis of the 49 loci identified from the GWAS of CRP on 46 disease endpoints or biomarker outcomes. The analyses generated evidence of associations of *FTO* (encoding FTO alpha-ketoglutarate dependent dioxygenase) with glucose, T2DM, CHD, heart failure and atrial fibrillation^45–47^; *IL6R* (encoding the interleukin-6 receptor) with stroke and CHD^17^; and *NLRP3* (encoding NLR family pyrin domain containing 3), *CD300LF* (encoding CMRF35-like molecule 1) and *IL1F10* (encoding IL-38) with white blood cell counts, in keeping with their role in the inflammatory response.^48,49^ However, we also noted evidence for a causal link between FDFT1 and ApoA1, triglycerides, T2DM, and systolic and diastolic blood pressure; and ZDHHC18 with several lipid measures.

**Fig. 3.**
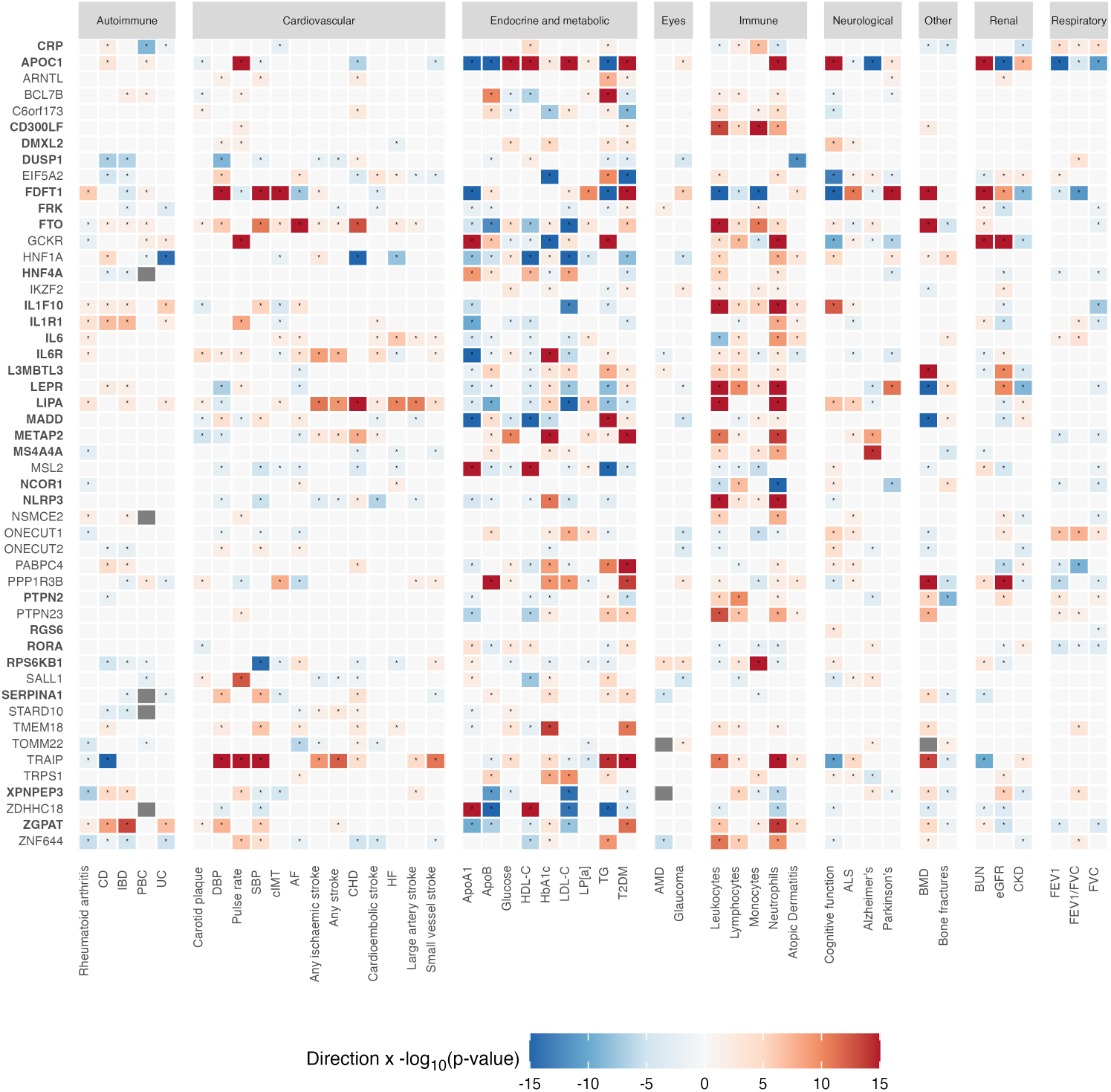
*trans*-weighted, *cis*-MR analysis of 49 genes implicated in a genome-wide association study of C-reactive protein for their direct causal effect on 46 end points. Cells are coloured according to the -log_10_ (p value) multiplied by the effect direction. Cells with an asterisk represent nominally significant estimates (p<0.05). The degree of significance is indicated by colour intensity, where red indicates higher values or risk of the outcome and blue for lower values or risk of the outcome; white and grey cells represent non-significant and indeterminate results respectively. Potentially druggable protein targets are highlighted in bold. Abbreviations: see **Supplementary Table 4** for phenotype abbreviations.

### Consistency between *trans*- and *cis*-weighted *cis*-MR analyses

We were able to check for consistency between *trans-weighted cis-MR* analysis using CRP-weighted estimates from Ligthart *et al* and the UKBB, and *cis*-weighted *cis*-MR analysis for six of 49 proteins assayed using the Somalogic platform from the deCODE proteomics GWAS,^50^ as well as 25 of 49 proteins using whole blood eQTL data from the Genotype-Tissue Expression database (GTEX)^51^ (**Fig. 4 and Extended Data Fig. 2**). Proteins were considered to demonstrate a direct causal effect on an outcome if a significant p value (p<0.0048; see Methods) was replicated for at least two of the four possible analyses. Likely causal associations were identified for IL6R with ApoA1 and monocyte count, and LEPR with neutrophil counts and BMD across all four analyses. A significant causal effect was observed for IL6R on most outcomes evaluated, particularly immune, autoimmune, cardiovascular, and endocrine and metabolic traits. For example, genetic variants at the *IL6R* locus, that are *trans*-for-CRP, showed a causal association with lower ischaemic stroke risk based on *cis*-MR analysis, both when *cis*-weighted by higher circulating s-IL6R concentration, which is an index of reduced signalling through the interleukin 6 receptor (OR 0.98, 95% CI 0.97, 0.98) and also when *trans*-weighted by higher circulating CRP concentration, an index of increased signalling through the interleukin 6 receptor (OR 1.39, 95% CI 1.24, 1.55; **Extended Data Fig. 3**).

**Fig. 4.**
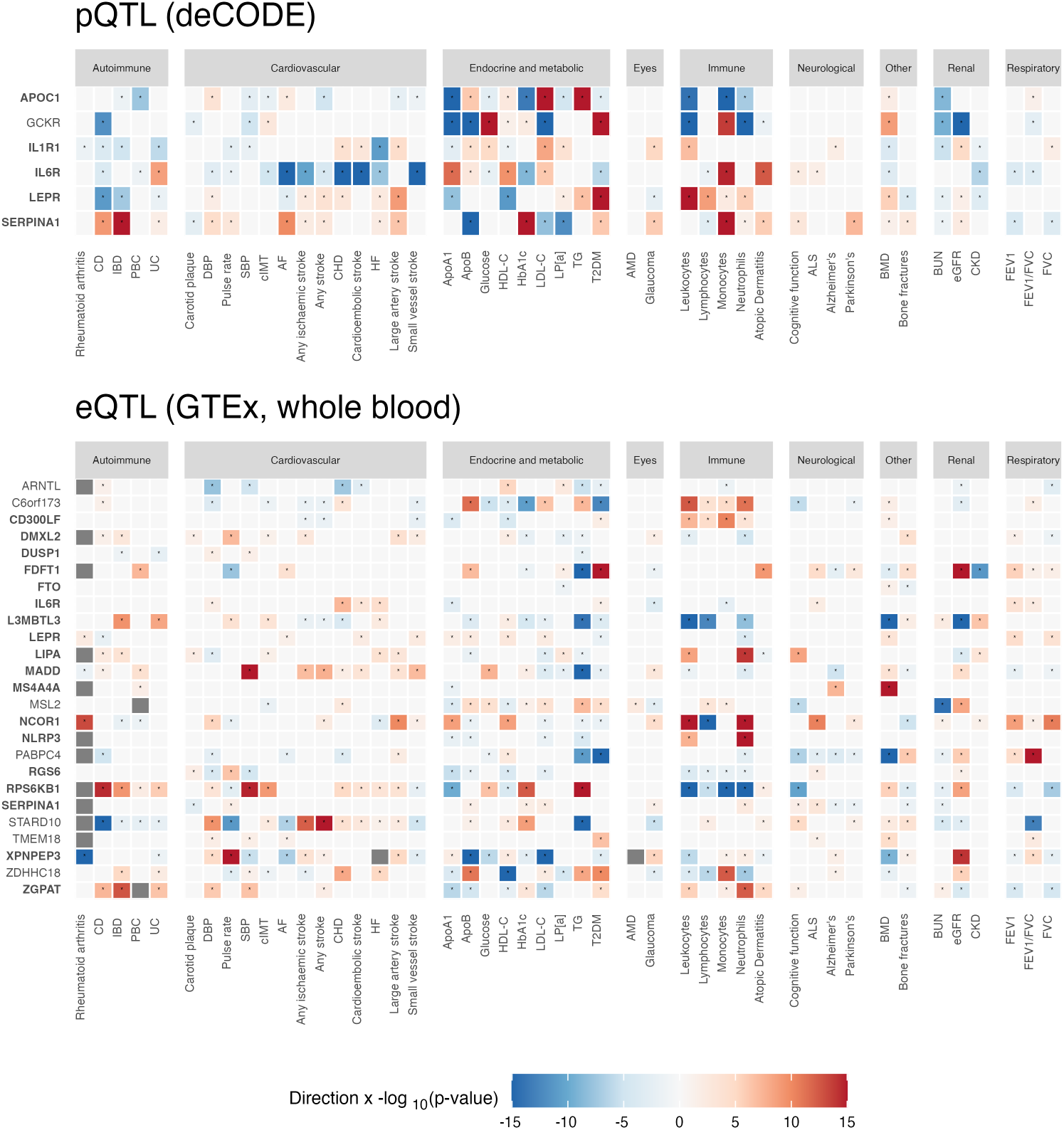
*cis*-weighted, *cis*-MR analysis of genes implicated in a genome-wide association study of C-reactive protein for their direct causal effect on 46 end points. Analyses are weighted by circulating protein concentration (pQTL; deCODE study) and encoded mRNA (eQTL; GTEx, whole blood). Cells are coloured according to the -log_10_ (p value) multiplied by the effect direction. Cells with an asterisk represent nominally significant estimates (p<0.05). The degree of significance is indicated by colour intensity, where red indicates higher values or risk of the outcome and blue for lower values or risk of the outcome; white and grey cells represent non-significant and indeterminate results respectively. Potentially druggable protein targets are highlighted in bold. Abbreviations: eQTL = expression quantitative trait loci; pQTL = protein quantitative trait loci; see **Supplementary Table 4** for phenotype abbreviations.

### Deconvoluting high-profile observational associations of CRP with common diseases

Associations of higher circulating CRP levels with several high-burden diseases have been consistently reported by non-genetic observational studies, including with T2DM, CHD, Alzheimer’s disease and age-related macular degeneration (AMD).^24,27,38,52^ Several MR analyses have followed, many of which have employed genome-wide rather than *cis*-MR analyses of CRP yielding inconsistent findings, as reviewed recently by Markozannes *et al*.^2^

For T2DM, we found that the *cis*-MR estimate for the causal effect of CRP itself on T2DM was not significant but that there was evidence from *trans*-weighted *cis*-MR analysis of a causal association of two CRP-associated proteins FTO and hepatocyte nuclear factor-4 alpha (HNF-4α), the product of *HNF4A* with T2DM (Fig. 4). The observational association between T2DM with CRP is therefore likely to be confounded by proteins such as FTO or HNF-4α, which are associated with both CRP and T2DM.

We also show that observational associations of CRP with CHD and Alzheimer’s disease are likely to be explained by other proteins that confound these associations, including lipase A, lysosomal acid type (encoded by *LIPA*) for CHD and apolipoprotein C1 (encoded by *APOC1*) and membrane spanning 4-domains A4A (encoded by *MS4A4A*) for Alzheimer’s disease (Fig. 4). Similarly, we found no evidence for a causal association between circulating CRP and age-related macular degeneration (AMD) using *cis*-MR analysis. However, *trans*-weighted *cis*-MR results suggest potentially causal roles for zinc-finger protein 644 (encoded by *ZNF644*), serpin family A member 1 (encoded by *SERPINA1*) and ribosomal protein S6 kinase B1 (encoded by *RPS6KB1*, Fig. 4).

### Potential therapeutic targets identified by MR-Fish

Of the 49 studied *trans*-for-CRP genes, 28 were predicted to encode potentially druggable proteins, of which 12 are targeted by existing licensed or unlicensed compounds (**Supplementary Table 2** and **Supplementary Table 3**). Some of the drug-target-indication relationships which were ‘rediscovered’ by applying *cis*-MR with *trans* weights include anakinra (IL1 receptor antagonist) for rheumatoid arthritis and lapaquistat (a squalene synthase inhibitor) for cholesterol lowering. Potential drug repurposing opportunities were also revealed in other cases, such as metreleptin (recombinant human leptin, which binds leptin receptor encoded by *LEPR*) for BMD, and alverine (HNF4A activator)^53^ and bisantrene (alpha-ketoglutarate-dependent dioxygenase FTO inhibitor) for T2DM.^54^

## Discussion

Using the circulating protein biomarker CRP as an example, we have shown that confounding in observational epidemiology and horizontal pleiotropy in MR analysis can be used to identify novel disease pathways and treatment targets. The MR Fish approach we illustrate here is a general one, applicable to any protein-disease association for which genetic variants are available that index the level of the protein of interest, some acting in *cis* and others in *trans*.

We selected CRP as an example to illustrate the approach because the assay of this protein is robust, there are numerous disease and biomarker associations of CRP identified in previous observational studies, and genetic instruments are available in *cis* and *trans*.^2^ Based on MR analysis using genetic variants located within or around the CRP gene (*cis*-MR), we found evidence of only eight causal associations, five with other biomarkers (neutrophil and monocyte counts, HDL-C concentration, FEV1, and FVC) and three with disease endpoints (CKD, PBC and bone fractures). The numerous observational associations of CRP, but few causal associations in *cis*-weighted *cis*-MR analysis is consistent with widespread confounding in non-genetic observational analysis, whereby a third factor associates with both CRP and the outcome of interest.

In observational epidemiology, much effort is spent on adjusting for rather than discovering confounders. Similarly, in the MR analysis of non-protein exposures (e.g. blood pressure or BMI) the selection of instruments from throughout the genome aims to balance the effects of horizontal pleiotropy (the hope being that positive effects at the locus are offset by negative effects at others). However, this cannot be relied on. Therefore, pleiotropy robust methods for MR analyses were developed (e.g. MR Egger) but these rely on further assumptions that may not be met.^55–58^ By considering proteins as a special category of exposure and using genetic instruments acting in *cis*, the risk of horizontal pleiotropy is substantially reduced, and *cis*-MR can be used reliably to separate causal from confounded associations.^13,15^ However, confounders in observational epidemiology and horizontal pleiotropy in MR analysis reflect causal disease pathways and could be regarded as of interest in biomedical research, rather than as a nuisance. Where such confounders are proteins, they could provide new therapeutic targets for the diseases concerned. However, until recently, there has been no systematic way to identify confounders in observational epidemiology. The approach exemplified here, which we term ‘MR-Fish’ for convenience, uses *cis*-MR analysis of the index or bait protein to interrogate confounding in the association between the bait and a disease. Where evident, it then harnesses the horizontal pleiotropy of genetic instruments that are *trans* for the index protein and uses them in *cis*-MR analyses of the proteins encoded by these genes. In so doing, the approach identifies other proteins (the ‘catch’), which are causal for the disease of interest, and which confound the association of the bait protein with disease.

For *cis*-MR analyses, we weighted the effect of the genetic instruments by the expression of the corresponding mRNA or the encoded protein itself (where such measures were available). We term this *cis*-weighted *cis*-MR analysis. For example, we were able to identify a causal association of genetic variants at the *IL6R* locus that are *trans*-for-CRP with a lower ischaemic stroke risk based on *cis*-MR analysis weighted by circulating s-IL6R concentration. Where measures of the expression of the corresponding mRNA or encoded protein are unavailable, or even when they are, it is possible to weight the effects of these genetic instruments by the original index protein (in this case CRP). In these analyses, which we term *trans*-weighted *cis*-MR, inference remains on the proteins encoded by the genes in which the instruments are located, rather than on CRP. For example, prior MR analysis of IL6R on ischaemic stroke weighted the effect of instruments at the *IL6R* locus by CRP but identified an effect on ischaemic stroke consistent with the *trans*-weighted *cis*-MR result identified here.^17^

A combination of either *cis*-weighted *cis*-MR or *trans*-weighted *cis*-MR analysis identified several other likely casual associations between CRP-associated proteins and a range of outcomes (**Fig. 3 – 5, Extended Data Fig. 2 & 3**). For example, we refuted a causal relationship of CRP on Alzheimer’s disease in a *cis*-MR analysis, despite previous reports of an observational association,^59^ which was suggested to be causal in a prior genome-wide MR analysis of CRP on Alzheimer’s disease (although not when excluding a single pleiotropic variant near the *APOE* gene).^60^ Instead, using *trans*-weighted *cis*-MR analysis, we found evidence for a causal association with Alzheimer’s disease for proteins encoded by genetic variants that are *trans* for CRP at the *APOE/C1* locus and the *MS4A4A* locus. The association of ApoE with Alzheimer’s disease is well established,^61^ and variants in the *MS4A*4A gene region are associated with soluble TREM2 concentrations in cerebrospinal fluid, which is associated with late-onset Alzheimer’s disease.^62–64^ Similarly, while our results suggested that CRP does not modulate AMD risk, there was evidence that various CRP-associated proteins may be causal for AMD, including those encoded at *ZNF644* and *SERPINA1*. Mutations in *ZNF644* are associated with high myopia which carries a high risk of myopic choroidal neovascularisation, a condition similar to neovascular AMD.^65^ Patients with high myopia may furthermore be at increased risk of developing neovascular AMD.^66^ *SERPINA1* encodes the serine protease inhibitor alpha-1-antitrypsin (AAT). Defects in this gene can cause alpha-1-antitrypsin disease, characterised by emphysema and liver disease (OMIM 613490).

**Fig. 5.**
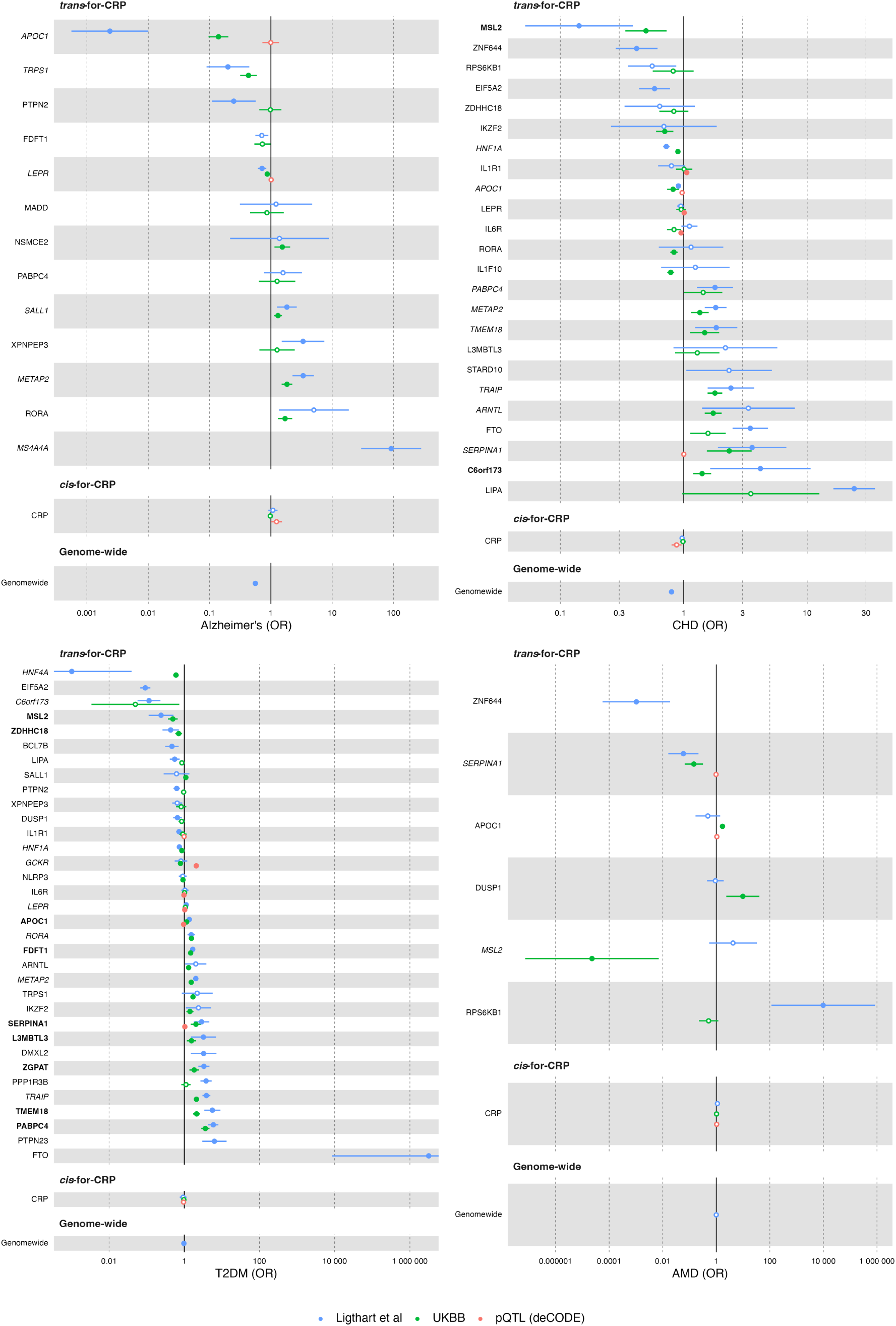
*cis*-Mendelian randomisation (MR) results with 95% confidence intervals from *trans*-weighted (by C-reactive protein (CRP); Ligthart et al, UK Biobank) and *cis*-weighted (protein quantitative trait loci) *cis*-MR analyses for selected outcomes. Genes are restricted to those with at least one significant estimate (p<0.0048) across the four analyses, ordered by effect size from the primary CRP-weighted analysis (Ligthart et al). Shaded circles and outlined circles indicate significant and non-significant results respectively. Italic genes: replicated in 2 analyses; bold genes: replicated in >2 analyses. Abbreviations: AMD = age-related macular degeneration; CHD = coronary heart disease; CRP = C-reactive protein; pQTL = protein quantitative trait loci; OR = odds ratio; T2DM = type 2 diabetes mellitus; UKBB = UK Biobank.

Interestingly, a proteomic study found upregulation of AAT in vitreous samples from eyes with AMD^67^ and AAT was shown to attenuate M1 microglia-mediated neuroinflammation in a mouse model of retinal degeneration.^68^ There is furthermore a well-described gene-environment interaction between *SERPINA1* and smoking,^69–71^ an important modifiable risk factor for AMD development and progression.^72^ Using *cis*-MR analysis of CRP, we also found evidence that the observational associations of CRP with T2DM are likely to be confounded. In this case, the genome-wide MR analysis also indicated a null effect because of balanced horizontal pleiotropy (Fig. 4). However, *trans* weighted *cis*-MR analysis using variants at the *FTO* and the *HNF4A* loci that are *trans* for CRP indicated a causal effect on T2DM of the proteins encoded by these genes. The effect of *FTO* variants on diabetes is likely through fat mass and insulin resistance. Inactivating mutations in *HNF4A* are a known cause of maturity onset diabetes of the young, the commonest monogenic form of diabetes. The associations we identified through *trans*-weighted *cis*-MR analysis of FDFT1 with ApoA1, triglycerides, T2DM, and systolic and diastolic blood pressure; and ZDHHC18 with several lipid measures are plausible because FDFT1 (also named squalene synthase) is the first specific enzyme in the mevalonate pathway involved in cholesterol biosynthesis,^73^ and ZDHHC18 is an enzyme in the Golgi apparatus that supports palmitoyl transferase activity. In *trans*-weighted *cis*-MR analysis there is the same degree of protection from horizontal pleiotropy as in *cis*-weighted *cis*-MR analysis. However, an important difference is that effects on outcomes in *trans*-weighted *cis*-MR analysis are expressed in terms of the effect on the bait protein (CRP in the current example), rather than in terms of the effect of the protein encoded by the gene in which the instruments reside. *trans*-weighted *cis*-MR analysis provides a valid test of the null hypothesis, but the effect estimates may be biased up or down depending on the relative effects of the genetic instruments used on the bait protein and, independently, on the disease (see **Mathematical appendix**).

This study has several strengths. We evaluated a broad range of CRP-outcome associations, while minimizing the risk of false discoveries by appropriately accounting for multiple testing. Nevertheless, the results from any MR study should not be interpreted in isolation, but rather considered in the context of pre-existing evidence. Compelling findings from a high-throughput MR analysis such as this may then serve as a useful tool to prioritize pursuit of new biological and therapeutic hypotheses.

The MR analyses drawing on transcriptomic and proteomic data were restricted to blood eQTL and pQTL data. This limits the ability to detect causal associations for certain proteins where expression is localised within specific cells and tissues. By excluding variants located outside the gene of interest, instruments for *cis*-MR may be underpowered in comparison to genome-wide MR. To address the issue of reduced power, we adopted a less stringent p-value threshold for *cis*-MR instrument selection with LD clumping of variants and modelling residual correlation using external reference data. This approach has previously been shown (analytically) to improve performance^13,74,75^ and any potential weak-instrument bias introduced by the inclusion of ‘null variants’ (i.e. variants that are not causally associated with the exposure) on average, attenuates results towards the null in two-sample MR.^76^ Using instruments in *cis* for MR analysis reduces but does not completely eliminate the risk of horizontal pleiotropy. The local correlation of genetic variants can undermine *cis*-MR analysis when a genetic variant employed as an instrument marks a nearby variant that influences disease risk through an adjacent gene and protein rather than the gene or protein of interest. Horizontal pleiotropy in *cis*-MR could also arise, in theory, if two adjacent genes were coregulated by the same genetic variant. In the current analysis, we ascribed effects to loci nominated by the authors of the CRP GWAS. However, the analytic pipeline we used incorporates assessment of horizontal pleiotropy arising due to linkage disequilibrium by identifying outlying variant associations with disease (that could indicate horizontal pleiotropy) when it favours use of pleiotropy-robust MR methods such as MR-Egger. A complementary approach is to use Bayesian statistical colocalization of genetic association signals with mRNA or protein expression of the encoding gene and the disease endpoint, although this approach is statistically conservative.^77^

Although we have exemplified the MR Fish approach using a single protein, in this case CRP, new technologies, population resources and analyses are providing the necessary data framework for conducting MR-Fish analysis at scale.^78^ Proteomics technologies from O-link and Somalogic now enable measurement of many thousands of proteins in stored plasma samples.^79,80^ The deployment of such technologies in large longitudinal cohort studies and national biobanks, enables investigation of the associations of thousands of proteins with hundreds of diseases at substantial scale.^81^ Many such associations are likely to be confounded, necessitating MR analysis of each protein to distinguish those which are causal. GWAS of the circulating proteome in the same population studies has already identified both *cis* and *trans* genetic variants for many such proteins, providing the necessary genetic instruments for *cis* analysis of each index protein, as well as MR Fish analysis.^50,82^ For example, over 2,900 proteins have been assayed using the O-link technology in samples from approximately 54,000 participants from UK Biobank. From this dataset 14,287 pQTL associations (for 2,414 proteins) were identified based on array-based genotyping of common variant, of which 1,955 were in *cis* and 12,332 were in *trans*.^83^ A recent analysis based on whole exome sequencing revealed 5,433 rare genotype-protein and 1,962 gene-protein associations. Together, these resources and others like them should enhance the genetic instruments available for MR-Fish analysis.

In summary, confounding in observational epidemiology and horizontal pleiotropy in MR analysis represent two aspects of the same phenomenon. Where proteins are the exposure of interest, genetic instruments acting in *cis* reduce the impact of horizontal pleiotropy in MR analysis compared to instruments which act in *trans*, as are incorporated in genome-wide MR analysis. However, instruments that are *trans* for an index protein can be utilized in a *cis*-MR analysis of the proteins encoded by the genes in which these variants reside. These analyses can not only help identify confounders of the association of the index protein with disease but, in so doing, generate new insights into disease pathogenesis and unveil new therapeutic targets.

## Methods

### Data sources

Genetic associations with CRP were obtained from a recent large meta-analysis of 88 studies (204,402 individuals) by Ligthart et al,^19^ which were associated through *cis*-MR to genetic associations with 46 traits (22 disease and 24 biomarker phenotypes; **Supplementary Table 4**). Replication data on protein quantitative trait loci (pQTL) and whole blood expression quantitative trait loci (eQTL) were leveraged from deCODE genetics and the Genotype-Tissue Expression (GTEx) project v8^51^ respectively.

### Instrument selection

The Ligthart et al. GWAS found 58 CRP-associated loci that passed genome-wide significance (p<5 x 10^-8^). These included *CRP* (ENSG00000132693), which is the *cis* acting gene for encoding CRP, and 57 *trans* loci. To showcase the difference between *cis* and genome-wide MR, instruments were selected from a 25kb flank around *CRP* (*cis*-weighted *cis*-MR) and compared to MR analyses sourcing genome-wide variants. Next, to illustrate how *cis*-MR can be utilised to deconvolute the genome-wide MR effects, which comprises the combined effects of CRP and many more *trans* loci, *cis*-MR was performed for 57 CRP-associated genetic loci from throughout the genome, *trans* weighting by CRP concentration (*trans*-weighted *cis*-MR), as assayed in the studies contributing to Ligthart et al. *Trans* loci for which MR analyses failed in >50% of outcomes at this step were removed, leaving 49 genes. Results were replicated utilising independent genetic effect estimates for CRP from the UKBB, as well as proteomics and transcriptomic data on the relevant protein directly encoded by these *trans*-CRP loci. For example, *cis*-MR was conducted to assess the causal relevance of IL6R to various outcomes utilising genetic effect estimates for CRP from Ligthart et al, estimates for CRP from the UKBB (both *trans*-weighted *cis*-MR), genetic effect estimates for IL6R plasma concentration, as well as using genetic estimates for IL6R mRNA expression (pQTL and eQTL *cis*-weighted *cis*-MR respectively). The CRP-weighted analyses selected instruments from within a 25kb region, while the pQTL and eQTL analyses used 200kb flanks. The larger flanking region used for the latter two analyses reflects increased confidence that the included variants are linked to the gene product, in comparison to the CRP-weighted analyses.^85^ There is furthermore some evidence that *trans*-eQTLs are often explained by *cis*-mediation.^86^

Genome-wide variants were selected using a 1 x 10^-8^ p value threshold and clumped to an r^2^ below 0.4. Given the smaller number of candidate variants, the *cis*-MR analyses included instruments based on a 1 x 10^-4^ p value filter. In both cases, local between-variant correlation (LD) was estimated from a random sample of 5000 UKBB participants of white British descent, after removing variants with a minor allele frequency below 0.01.

### Mendelian randomisation analyses

MR analyses were conducted using a generalised least squares (GLS) implementation of the inverse variance weighted (IVW) estimator, and the horizontal pleiotropy robust MR-Egger.^74^ In each case, residual horizontal pleiotropy was accounted for utilising the aforementioned UKBB LD reference sample. Genome-wide and *cis*-MR were compared using the IVW model and without removing overinfluential variants. For all other analyses, a model selection algorithm was applied to determine which of these estimators had most data.^13^ To further limit the potential of horizontal pleiotropy potentially overinfluential variants were removed, i.e., those with a high leverage statistic or with a relatively large contribution to the heterogeneity statistic.

### Identifying potential therapeutic targets

The OpenTargets^87^ API was queried to identify druggable CRP-associated proteins and which of these are targeted by existing compounds. Protein targets were deemed potentially druggable according to the following criteria: (i) the target has clinical precedence with Phase I, II, III or IV drugs; (ii) the target has a binding site suitable for small molecule binding (high-quality ligand, high-quality pocket or considered druggable as per Finan et al.’s Druggable Genome Pipeline^88^); (iii) the target has an accessible epitope for antibody based therapy with high confidence from either Uniprot^89^ or Gene Ontolology^90^ that the subcellular location of the target is either plasma membrane, extracellular region/matrix, or secretion.

### Statistical analysis

Point estimates are presented as odds ratios or mean differences for outcomes (units are detailed in **Supplementary Table 4**) per unit change in CRP (log(mg/L) for Ligthart et al, mg/L for UKBB, and SD expression or circulating levels for transcriptomics and proteomics data; **Supplementary Table 5**) and 95% confidence intervals. Difference between genome-wide and *cis*-MR estimates were tested using formal interaction tests at a Bonferroni-corrected p value threshold of 0.001 (0.05/46 outcomes).^91^ *cis*-MR results (both *cis*-weighted and *trans*-weighted) were considered statistically significant for protein-outcome pairs with at least two replicated signals at a multiplicity-corrected p value threshold of 0.0048 (sqrt(0.05/(46 outcomes x 49 proteins))). For the empirical validation of applying *cis*-MR with *trans* weights (*trans*-weighted *cis*-MR; **Mathematical appendix**), the proportion of predicted effect directions matching those observed was tested against a null hypothesis of 50% (random chance) using Clopper-Pearson confidence intervals for a proportion.^92^ Analyses were conducted using Python v3.7.4 (for GNU Linux),^93^ Pandas v0.25,^94^ Numpy v1.15,^95^ R v4.1.0 (for GNU macOS),^96^ ggforestplot,^97^ targets,^100^ tarchetypes,^99^ tidyverse,^100^ workflowr,^101^ flextable,^102^ gtsummary^103^ and knitr.^104^

## Supporting information

Mathematical appendix

## Data Availability

All data produced in the present work are contained in the manuscript.

## Acknowledgements

This research was supported by the UCL NIHR Biomedical Research Centre, the UCL British Heart Foundation Research Accelerator, the UKRI-NIHR funded Multimorbidity Mechanisms and Therapeutics Research Collaborative (MR/V033867/1), and the National Institute for Health and Care Research (NIHR) Biomedical Research Centre based at Moorfields Eye Hospital NHS Foundation Trust and UCL Institute of Ophthalmology. The views expressed are those of the author(s) and not necessarily those of the NHS, the NIHR or the Department of Health and Social Care.

ANW is supported by the Wellcome Trust (220558/Z/20/Z; 224390/Z/21/Z).

KVS is supported by grants from Fight for Sight (1956A) and The Desmond Foundation.

APK is supported by a UK Research and Innovation Future Leaders Fellowship, an Alcon Research Institute Young Investigator Award and a Lister Institute of Preventive Medicine Award. For the purpose of open access, the author has applied a Creative Commons Attribution (CC BY) licence to any Author Accepted Manuscript version arising.

CE, and AT received a proportion of their financial support from the UK Department of Health through an award made by the National Institute for Health Research to Moorfields Eye Hospital NHS Foundation Trust and UCL Institute of Ophthalmology for a Biomedical Research Centre for Ophthalmology. CE receives a proportion of financial support to Moorfields Eye Hospital NHS Foundation Trust from the Lowy Medical Research Institute.

AFS is supported by BHF grant PG/22/10989, the UCL BHF Research Accelerator AA/18/6/34223, and the National Institute for Health and Care Research University College London Hospitals Biomedical Research Centre. This work was funded by UK Research and Innovation (UKRI) under the UK government’s Horizon Europe funding guarantee EP/Z000211/1.

ADH is an NIHR Senior Investigator.

## Data availability

All MR results are provided in **Supplementary Table 5**.

## Extended Data Figure Captions

**Extended Data Fig. 1.**
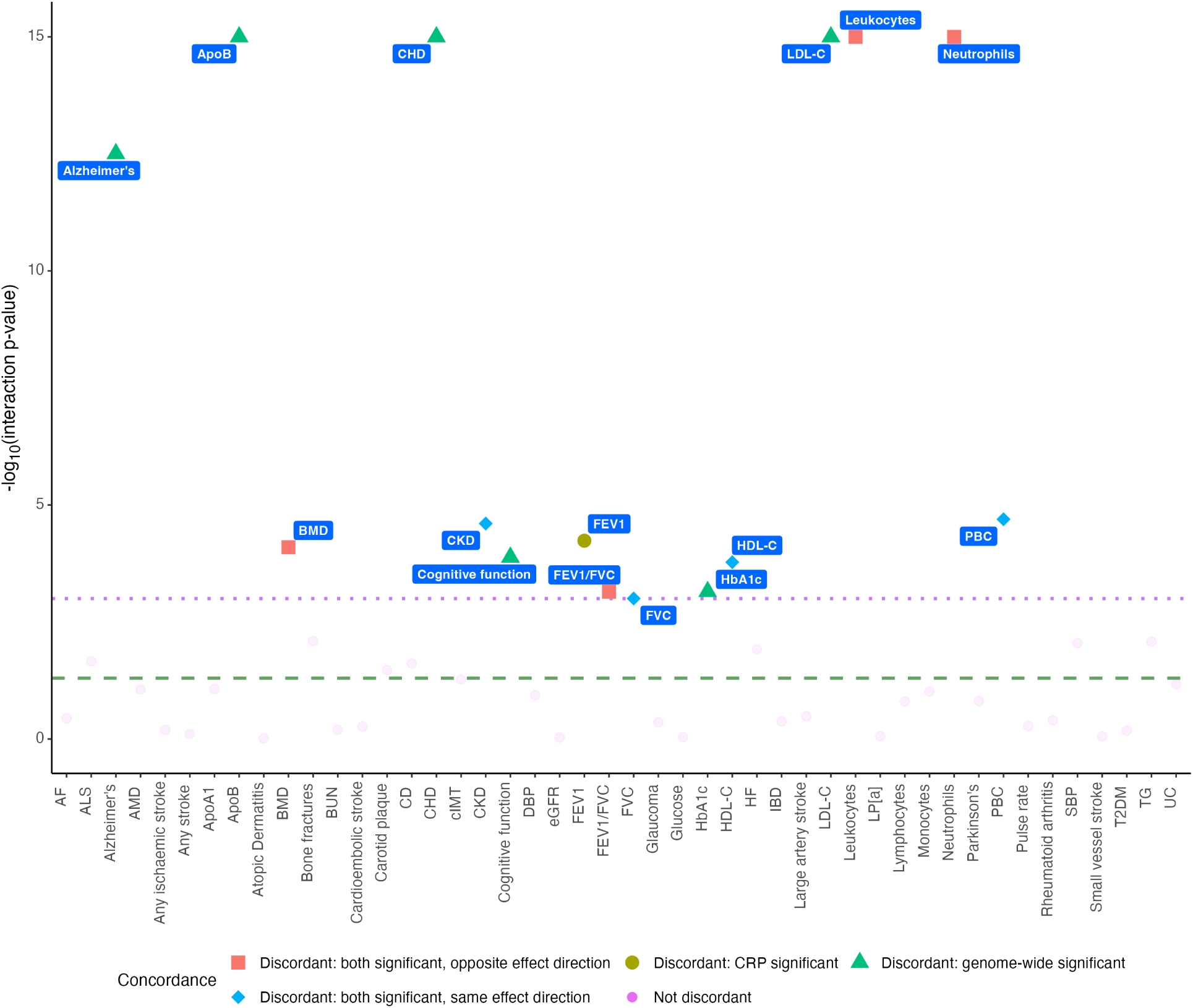
Genome-wide and *cis*-Mendelian randomisation estimates for the causal relevance of C-reactive protein to all outcomes were compared by calculating interaction p-values (Wald test), transformed here by -log10. The dashed green and dotted purple lines correspond to interaction p-values of 0.05 and 0.001 respectively. Abbreviations: see **Supplementary Table 4** for phenotype abbreviations.

**Extended Data Fig. 2.**
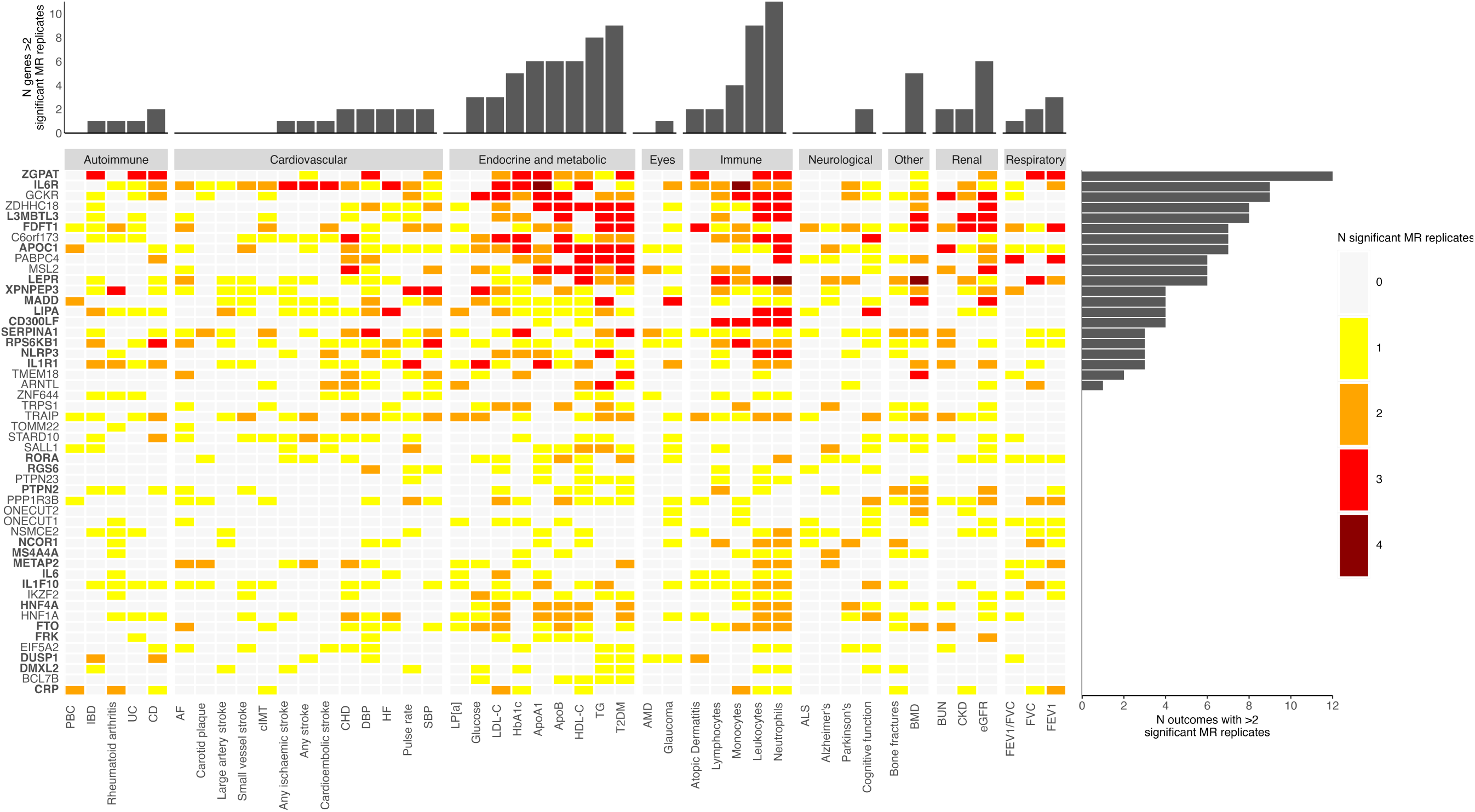
Heatmap highlighting gene-outcome pairs with significant *cis*-Mendelian randomisation results (p<0.0048) across multiple replication analyses (exposure summary statistics: C-reactive protein (CRP) Ligthart et al, CRP UK Biobank, deCODE genetics protein quantitative trait loci, Genotype-Tissue Expression Project expression quantitative trait loci). Marginal bar charts indicate the number of genes or outcomes with ≥3 significant replicates per outcome or gene respectively. Potentially druggable targets are highlighted in bold. Abbreviations: MR = Mendelian randomization; see **Supplementary Table 4** for phenotype abbreviations.

**Extended Data Fig. 3.**
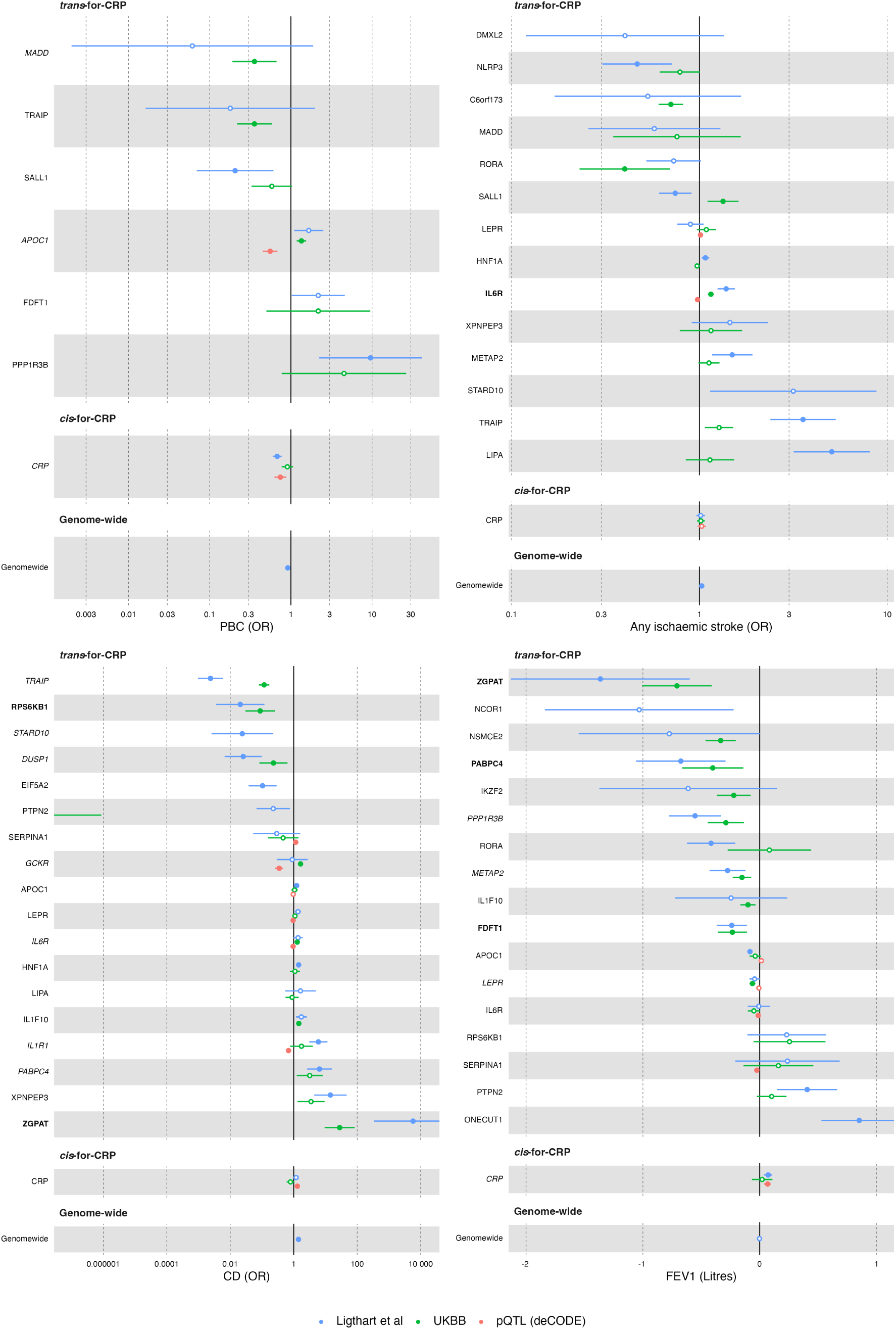
*cis*-Mendelian randomisation (MR) results with 95% confidence intervals from *trans*-weighted (by C-reative protein (CRP); Ligthart et al, UK Biobank) and *cis*-weighted (protein quantitative trait loci) *cis*-MR analyses for selected outcomes. Genes are restricted to those with at least one significant estimate (p<0.0048) across the four analyses, ordered by effect size from the primary CRP-weighted analysis (Ligthart et al). Shaded circles and outlined circles indicate significant and non-significant results respectively. Italic genes: replicated in 2 analyses; bold genes: replicated in >2 analyses. Abbreviations: CD = Crohn’s disease; CRP = C-reactive protein; FEV1 = forced expiratory volume in one second; pQTL = protein quantitative trait loci; OR = odds ratio; PBC = primary biliary cirrhosis; UKBB = UK Biobank.

## References

1. Biomarkers Definitions Working Group. Biomarkers and surrogate endpoints: preferred definitions and conceptual framework. Clin. Pharmacol. Ther. 69, 89–95 (2001).

2. Markozannes, G. et al. Global assessment of C-reactive protein and health-related outcomes: an umbrella review of evidence from observational studies and Mendelian randomization studies. Eur. J. Epidemiol. 36, 11–36 (2021).

3. Lawlor, D. A., Davey Smith, G., Kundu, D., Bruckdorfer, K. R. & Ebrahim, S. Those confounded vitamins: what can we learn from the differences between observational versus randomised trial evidence? Lancet Lond. Engl. 363, 1724–1727 (2004).

4. Smith, G. D. et al. Clustered environments and randomized genes: a fundamental distinction between conventional and genetic epidemiology. PLoS Med. 4, e352 (2007).

5. Pepys, M. B. The Pentraxins 1975-2018: Serendipity, Diagnostics and Drugs. Front. Immunol. 9, 2382 (2018).

6. Van Norman, G. A. Drugs, Devices, and the FDA: Part 1: An Overview of Approval Processes for Drugs. JACC Basic Transl. Sci. 1, 170–179 (2016).

7. Arrowsmith, J. & Miller, P. Trial watch: phase II and phase III attrition rates 2011-2012. Nat. Rev. Drug Discov. 12, 569 (2013).

8. Schmidt, A. F., Hingorani, A. D. & Finan, C. Human Genomics and Drug Development. Cold Spring Harb. Perspect. Med. a039230 (2021) doi:10.1101/cshperspect.a039230.

9. Davies, N. M., Holmes, M. V. & Smith, G. D. Reading Mendelian randomisation studies: a guide, glossary, and checklist for clinicians. BMJ 362, k601 (2018).

10. Smith, G. D. & Ebrahim, S. Mendelian randomization: prospects, potentials, and limitations. Int. J. Epidemiol. 33, 30–42 (2004).

11. Smith, G. D. & Ebrahim, S. ‘Mendelian randomization’: can genetic epidemiology contribute to understanding environmental determinants of disease? Int. J. Epidemiol. 32, 1–22 (2003).

12. Hingorani, A. & Humphries, S. Nature’s randomised trials. Lancet Lond. Engl. 366, 1906–1908 (2005).

13. Schmidt, A. F. et al. Genetic drug target validation using Mendelian randomisation. Nat. Commun. 11, 3255 (2020).

14. Dehghan, A. et al. Meta-analysis of genome-wide association studies in >80 000 subjects identifies multiple loci for C-reactive protein levels. Circulation 123, 731–738 (2011).

15. Swerdlow, D. I. et al. Selecting instruments for Mendelian randomization in the wake of genome-wide association studies. Int. J. Epidemiol. 45, 1600–1616 (2016).

16. Lawlor, D. A., Harbord, R. M., Sterne, J. A. C., Timpson, N. & Davey Smith, G. Mendelian randomization: using genes as instruments for making causal inferences in epidemiology. Stat. Med. 27, 1133–1163 (2008).

17. Georgakis, M. K. et al. Interleukin-6 Signaling Effects on Ischemic Stroke and Other Cardiovascular Outcomes: A Mendelian Randomization Study. Circ. Genomic Precis. Med. 13, e002872 (2020).

18. Eiriksdottir, G. et al. Association between C reactive protein and coronary heart disease: Mendelian randomisation analysis based on individual participant data. Bmj 342, 425 (2011).

19. Ligthart, S. et al. Genome Analyses of >200,000 Individuals Identify 58 Loci for Chronic Inflammation and Highlight Pathways that Link Inflammation and Complex Disorders. Am. J. Hum. Genet. 103, 691–706 (2018).

20. Said, S. et al. Genetic analysis of over half a million people characterises C-reactive protein loci. Nat. Commun. 13, 2198 (2022).

21. Xu, W. et al. The relationship between high-sensitivity C-reactive protein and ApoB, ApoB/ApoA1 ratio in general population of China. Endocrine 42, 132–138 (2012).

22. de Pablo, P., Cooper, M. S. & Buckley, C. D. Association between bone mineral density and C-reactive protein in a large population-based sample. Arthritis Rheum. 64, 2624– 2631 (2012).

23. Xu, R., Zhang, Y., Gao, X., Wan, Y. & Fan, Z. High-Sensitivity CRP (C-Reactive Protein) Is Associated With Incident Carotid Artery Plaque in Chinese Aged Adults. Stroke 50, 1655–1660 (2019).

24. Noble, J. M. et al. Association of C-reactive protein with cognitive impairment. Arch. Neurol. 67, 87–92 (2010).

25. Sesso, H. D. et al. C-reactive protein and the risk of developing hypertension. JAMA 290, 2945–2951 (2003).

26. Ahmadi-Abhari, S., Kaptoge, S., Luben, R. N., Wareham, N. J. & Khaw, K.-T. Longitudinal association of C-reactive protein and lung function over 13 years: The EPIC-Norfolk study. Am. J. Epidemiol. 179, 48–56 (2014).

27. Aronson, D. et al. Association between fasting glucose and C-reactive protein in middle-aged subjects. Diabet. Med. J. Br. Diabet. Assoc. 21, 39–44 (2004).

28. Kim, K.-I. et al. CRP level and HDL cholesterol concentration jointly predict mortality in a Korean population. Am. J. Med. 125, 787–795.e4 (2012).

29. Dongway, A. C., Faggad, A. S., Zaki, H. Y. & Abdalla, B. E. C-reactive protein is associated with low-density lipoprotein cholesterol and obesity in type 2 diabetic Sudanese. Diabetes Metab. Syndr. Obes. Targets Ther. 8, 427–435 (2015).

30. Puri, R. et al. Effect of C-Reactive Protein on Lipoprotein(a)-Associated Cardiovascular Risk in Optimally Treated Patients With High-Risk Vascular Disease: A Prespecified Secondary Analysis of the ACCELERATE Trial. JAMA Cardiol. 5, 1136–1143 (2020).

31. Sproston, N. R. & Ashworth, J. J. Role of C-Reactive Protein at Sites of Inflammation and Infection. Front. Immunol. 9, 754 (2018).

32. Whelton, S. P. et al. Association between resting heart rate and inflammatory biomarkers (high-sensitivity C-reactive protein, interleukin-6, and fibrinogen) (from the Multi-Ethnic Study of Atherosclerosis). Am. J. Cardiol. 113, 644–649 (2014).

33. Firdous, S. Correlation of CRP, fasting serum triglycerides and obesity as cardiovascular risk factors. J. Coll. Physicians Surg.--Pak. JCPSP 24, 308–313 (2014).

34. Amer, M. S. et al. Association of high-sensitivity C-reactive protein with carotid artery intima-media thickness in hypertensive older adults. J. Am. Soc. Hypertens. JASH 5, 395– 400 (2011).

35. Kugler, E. et al. C reactive protein and long-term risk for chronic kidney disease: a historical prospective study. J. Nephrol. 28, 321–327 (2015).

36. Galea, R. et al. Inflammation and C-reactive protein in atrial fibrillation: cause or effect? Tex. Heart Inst. J. 41, 461–468 (2014).

37. Keizman, D. et al. Low-grade systemic inflammation in patients with amyotrophic lateral sclerosis. Acta Neurol. Scand. 119, 383–389 (2009).

38. Seddon, J. M., Gensler, G., Milton, R. C., Klein, M. L. & Rifai, N. Association between C-reactive protein and age-related macular degeneration. JAMA 291, 704–710 (2004).

39. Shadick, N. A. et al. C-reactive protein in the prediction of rheumatoid arthritis in women. Arch. Intern. Med. 166, 2490–2494 (2006).

40. Vekaria, A. S. et al. Moderate-to-severe atopic dermatitis patients show increases in serum C-reactive protein levels, correlating with skin disease activity. F1000Research 6, 1712 (2017).

41. Henriksen, M. et al. C-reactive protein: a predictive factor and marker of inflammation in inflammatory bowel disease. Results from a prospective population-based study. Gut 57, 1518–1523 (2008).

42. Leibovitch, I. et al. C-reactive protein levels in normal tension glaucoma. J. Glaucoma 14, 384–386 (2005).

43. Cash, W. J. et al. Primary biliary cirrhosis is associated with oxidative stress and endothelial dysfunction but not increased cardiovascular risk. Hepatol. Res. Off. J. Jpn. Soc. Hepatol. 40, 1098–1106 (2010).

44. Qiu, X. et al. C-Reactive Protein and Risk of Parkinson’s Disease: A Systematic Review and Meta-Analysis. Front. Neurol. 10, 384 (2019).

45. Gustavsson, J. et al. FTO genotype, physical activity, and coronary heart disease risk in Swedish men and women. Circ. Cardiovasc. Genet. 7, 171–177 (2014).

46. Frayling, T. M. et al. A common variant in the FTO gene is associated with body mass index and predisposes to childhood and adult obesity. Science 316, 889–894 (2007).

47. Scott, L. J. et al. A genome-wide association study of type 2 diabetes in Finns detects multiple susceptibility variants. Science 316, 1341–1345 (2007).

48. Swanson, K. V., Deng, M. & Ting, J. P.-Y. The NLRP3 inflammasome: molecular activation and regulation to therapeutics. Nat. Rev. Immunol. 19, 477–489 (2019).

49. Diaz-Barreiro, A., Huard, A. & Palmer, G. Multifaceted roles of IL-38 in inflammation and cancer. Cytokine 151, 155808 (2022).

50. Ferkingstad, E. et al. Large-scale integration of the plasma proteome with genetics and disease. Nat. Genet. 53, 1712–1721 (2021).

51. GTEx Consortium et al. Genetic effects on gene expression across human tissues. Nature 550, 204–213 (2017).

52. Ridker, P. M., Rifai, N., Rose, L., Buring, J. E. & Cook, N. R. Comparison of C-reactive protein and low-density lipoprotein cholesterol levels in the prediction of first cardiovascular events. N. Engl. J. Med. 347, 1557–1565 (2002).

53. Lee, S.-H., Athavankar, S., Cohen, T., Kiselyuk, A. & Levine, F. Reversal of Lipotoxic Effects on the Insulin Promoter by Alverine and Benfluorex: Identification as HNF4α Activators. ACS Chem. Biol. 8, 1730–1736 (2013).

54. Bornaque, F. et al. Glucose Regulates m6A Methylation of RNA in Pancreatic Islets. Cells 11, 291 (2022).

55. Bowden, J., Davey Smith, G. & Burgess, S. Mendelian randomization with invalid instruments: effect estimation and bias detection through Egger regression. Int. J. Epidemiol. 44, 512–525 (2015).

56. Bowden, J., Davey Smith, G., Haycock, P. C. & Burgess, S. Consistent Estimation in Mendelian Randomization with Some Invalid Instruments Using a Weighted Median Estimator. Genet. Epidemiol. 40, 304–314 (2016).

57. Verbanck, M., Chen, C.-Y., Neale, B. & Do, R. Detection of widespread horizontal pleiotropy in causal relationships inferred from Mendelian randomization between complex traits and diseases. Nat. Genet. 50, 693–698 (2018).

58. Foley, C. N., Mason, A. M., Kirk, P. D. W. & Burgess, S. MR-Clust: clustering of genetic variants in Mendelian randomization with similar causal estimates. Bioinforma. Oxf. Engl. 37, 531–541 (2021).

59. Koyama, A. et al. The role of peripheral inflammatory markers in dementia and Alzheimer’s disease: a meta-analysis. J. Gerontol. A. Biol. Sci. Med. Sci. 68, 433–440 (2013).

60. Larsson, S. C., et al. Modifiable pathways in Alzheimer’s disease: Mendelian randomisation analysis. The BMJ 359, j5375 (2017).

61. Serrano-Pozo, A., Das, S. & Hyman, B. T. APOE and Alzheimer’s disease: advances in genetics, pathophysiology, and therapeutic approaches. Lancet Neurol. 20, 68–80 (2021).

62. Deming, Y. et al. The MS4A gene cluster is a key modulator of soluble TREM2 and Alzheimer’s disease risk. Sci. Transl. Med. 11, (2019).

63. You, S.-F. et al. MS4A4A modifies the risk of Alzheimer disease by regulating lipid metabolism and immune response in a unique microglia state. MedRxiv Prepr. Serv. Health Sci. 2023.02.06.23285545 (2023) doi:10.1101/2023.02.06.23285545.

64. Ulland, T. K. & Colonna, M. TREM2 - a key player in microglial biology and Alzheimer disease. Nat. Rev. Neurol. 14, 667–675 (2018).

65. Shi, Y. et al. Exome sequencing identifies ZNF644 mutations in high myopia. PLoS Genet. 7, e1002084 (2011).

66. Corbelli, E. et al. Prevalence and Phenotypes of Age-Related Macular Degeneration in Eyes With High Myopia. Invest. Ophthalmol. Vis. Sci. 60, 1394–1402 (2019).

67. Koss, M. J. et al. Proteomics of vitreous humor of patients with exudative age-related macular degeneration. PloS One 9, e96895 (2014).

68. Zhou, T. et al. Alpha-1 Antitrypsin Attenuates M1 Microglia-Mediated Neuroinflammation in Retinal Degeneration. Front. Immunol. 9, 1202 (2018).

69. Deng, X., Yuan, C.-H. & Chang, D. Interactions between single nucleotide polymorphism of SERPINA1 gene and smoking in association with COPD: a case-control study. Int. J. Chron. Obstruct. Pulmon. Dis. 12, 259–265 (2017).

70. Silverman, E. K. et al. Genetic epidemiology of severe, early-onset chronic obstructive pulmonary disease. Risk to relatives for airflow obstruction and chronic bronchitis. Am. J. Respir. Crit. Care Med. 157, 1770–1778 (1998).

71. Kim, W. et al. Genome-Wide Gene-by-Smoking Interaction Study of Chronic Obstructive Pulmonary Disease. Am. J. Epidemiol. 190, 875–885 (2021).

72. Velilla, S. et al. Smoking and age-related macular degeneration: review and update. J. Ophthalmol. 2013, 895147 (2013).

73. Trapani, L., Segatto, M., Ascenzi, P. & Pallottini, V. Potential role of nonstatin cholesterol lowering agents. IUBMB Life 63, 964–971 (2011).

74. Burgess, S., Zuber, V., Valdes-Marquez, E., Sun, B. B. & Hopewell, J. C. Mendelian randomization with fine-mapped genetic data: Choosing from large numbers of correlated instrumental variables. Genet. Epidemiol. 41, 714–725 (2017).

75. Dudbridge, F. Power and Predictive Accuracy of Polygenic Risk Scores. PLOS Genet. 9, e1003348 (2013).

76. Burgess, S., Thompson, S. G., & CRP CHD Genetics Collaboration. Avoiding bias from weak instruments in Mendelian randomization studies. Int. J. Epidemiol. 40, 755–764 (2011).

77. Giambartolomei, C. et al. Bayesian test for colocalisation between pairs of genetic association studies using summary statistics. PLoS Genet. 10, e1004383 (2014).

78. Suhre, K., McCarthy, M. I. & Schwenk, J. M. Genetics meets proteomics: perspectives for large population-based studies. Nat. Rev. Genet. 22, 19–37 (2021).

79. Olink. Olink https://olink.com/.

80. SomaLogic. SomaLogic https://somalogic.com/.

81. Carrasco-Zanini, J. et al. Proteomic prediction of common and rare diseases. 2023.07.18.23292811 Preprint at 10.1101/2023.07.18.23292811 (2023).

82. Sun, B. B. et al. Genomic atlas of the human plasma proteome. Nature 558, 73–79 (2018).

83. Sun, B. B. et al. Plasma proteomic associations with genetics and health in the UK Biobank. Nature 622, 329–338 (2023).

84. Dhindsa, R. S. et al. Rare variant associations with plasma protein levels in the UK Biobank. Nature 622, 339–347 (2023).

85. Fauman, E. B. & Hyde, C. An optimal variant to gene distance window derived from an empirical definition of cis and trans protein QTLs. BMC Bioinformatics 23, 169 (2022).

86. Pierce, B. L. et al. Mediation Analysis Demonstrates That Trans-eQTLs Are Often Explained by Cis-Mediation: A Genome-Wide Analysis among 1,800 South Asians. PLOS Genet. 10, e1004818 (2014).

87. Ochoa, D. et al. The next-generation Open Targets Platform: reimagined, redesigned, rebuilt. Nucleic Acids Res. 51, D1353–D1359 (2023).

88. Finan, C. et al. The druggable genome and support for target identification and validation in drug development. Sci. Transl. Med. 9, eaag1166 (2017).

89. UniProt. UniProt https://www.uniprot.org/.

90. Gene Ontology Resource. Gene Ontology Resource http://geneontology.org/.

91. Schmidt, A. F. et al. Exploring interaction effects in small samples increases rates of false-positive and false-negative findings: results from a systematic review and simulation study. J. Clin. Epidemiol. 67, 821–829 (2014).

92. Clopper, C. J. & Pearson, E. S. THE USE OF CONFIDENCE OR FIDUCIAL LIMITS ILLUSTRATED IN THE CASE OF THE BINOMIAL. Biometrika 26, 404–413 (1934).

93. Van Rossum, G. & Drake Jr, F. L. Python Reference Manual. (Centrum voor Wiskunde en Informatica Amsterdam, 1995).

94. Wes McKinney. Data Structures for Statistical Computing in Python. in Proceedings of the 9th Python in Science Conference (eds. van der Walt, S. & Jarrod Millman) 56–61 (2010). doi:10.25080/Majora-92bf1922-00a.

95. Harris, C. R. et al. Array programming with NumPy. Nature 585, 357–362 (2020).

96. R Core Team. R: A Language and Environment for Statistical Computing. https://www.R-project.org/ (2022).

97. Scheinin, I. et al. Ggforestplot: Forestplots of Measures of Effects and Their Confidence Intervals. (2022).

98. Landau, W. M. The targets R package: a dynamic Make-like function-oriented pipeline toolkit for reproducibility and high-performance computing. J. Open Source Softw. 6, 2959 (2021).

99. Landau, W. M. Tarchetypes: Archetypes for Targets. (2021).

100. Wickham, H. et al. Welcome to the tidyverse. J. Open Source Softw. 4, 1686 (2019).

101. Blischak, J. D., Carbonetto, P. & Stephens, M. Creating and sharing reproducible research code the workflowr way [version 1; peer review: 3 approved]. F1000Research 8, (2019).

102. Gohel, D. Flextable: Functions for Tabular Reporting. https://CRAN.R-project.org/package=flextable (2022).

103. Sjoberg, D. D., Whiting, K., Curry, M., Lavery, J. A. & Larmarange, J. Reproducible summary tables with the gtsummary package. R J. 13, 570–580 (2021).

104. Xie, Y. Knitr: A General-Purpose Package for Dynamic Report Generation in R. https://yihui.org/knitr/ (2022).

