## Supplementary material for "Harnessing confounding and genetic pleiotropy to identify causes of disease through proteomics and Mendelian randomisation – ‘MR Fish’": Mathematical appendix

This appendix provides mathematical derivations for the effect estimates of the various types of Mendelian randomisation (MR) analysis (*cis*-MR, *trans*-MR, genome-wide MR and *trans*-weighted *cis*-MR). For each, we indicate potential sources of pleiotropy (see **Figure 2, Mathematical appendix Table A1**).

### *cis*-MR analysis

In **Figure 2a**, unmeasured or unknown variables ($U$) confound the directly observed association of CRP with a disease ($D$). MR analysis may be used to address confounding by selecting as an instrumental variable, genetic instruments associated with CRP. In **Figure 2b**, the instruments are within or in the vicinity of the CRP gene and act in *cis*. The MR estimate of the causal effect of CRP on $D$ in this case is given by $\frac{(\sigma_{CRP} \cdot \delta_D^{CRP}) + \emptyset_D^{\mathrm{GCRP}}}{\sigma_{CRP}}$ (**Mathematical appendix, Table A1**). Here the term $\sigma$ refers to the effect of variants in a gene on the encoded protein acting in *cis* (e.g., $\sigma_{CRP}$ refers to the effect of variants in the $CRP$ gene on CRP concentration); $\delta$ to the direct effect of a protein on disease (e.g., $\delta_D^{CRP}$ refers to the effect of CRP on disease $D$); and $\emptyset$ to pathways leading to horizontal pleiotropy (e.g., $\emptyset_D^{\mathrm{GCRP}}$ refers to horizontal pleiotropy between variants in the gene encoding CRP and disease $D$, which might arise, for example, due to linkage disequilibrium with variants in a nearby gene). If the assumptions of MR analysis are fully met, the pleiotropy term $\emptyset_D^{\mathrm{GCRP}} = 0$ and the Mendelian randomisation estimate is equal to the causal effect of CRP on D ( $\delta_D^{CRP}$).

*trans- and genome-wide MR analysis*

Genetic variants outside the $CRP$ gene (acting in *trans*) might be considered as potential instruments in a MR analysis of $CRP$ on $D$ (**Figure 2c**), e.g., variants in a gene $G_1$ encoding a protein $P_1$ which has a downstream effect on CRP. However, an analysis of this type runs a higher risk of violating the no horizontal pleiotropy assumption because there may be several causal pathways from $G_1$ to $D$, some independent of CRP. The MR estimate of the causal effect of CRP on $D$ in this case is given by $\frac{(\sigma_{P1} \cdot \delta_D^{P1}) + (\sigma_{P1} \cdot \delta_{CRP}^{P1} . \delta_D^{CRP} + \emptyset_D^{G1}) + (\emptyset_{CRP}^{\mathrm{G1}} . \delta_D^{CRP})}{(\sigma_{P1} \cdot \delta_{CRP}^{P1}) + \emptyset_{CRP}^{\mathrm{G1}}}$ (see **Figure 2** and **Mathematical appendix, Table A1**). If $\emptyset_D^{G1} = \emptyset_{CRP}^{\mathrm{G1}} = 0$, the MR estimate reduces to $\frac{(\delta_D^{P1} + \delta_{CRP}^{P1}) \cdot \delta_D^{CRP}}{\delta_{CRP}^{P1}}$. Only if $\delta_D^{P1} = 0$ (i.e., no causal effect of $P_1$ on $D$) does the MR estimate equate to $\delta_D^{CRP}$ , the causal effect of CRP on $D$. In genome-wide MR analysis, a single genetic instrument is selected from multiple loci throughout the genome. Although this may include a variant within the CRP gene (acting in *cis*), the majority are from outside the CRP gene (acting in *trans*) and, for this reason, genome-wide MR analysis of CRP may be more prone to generating a biased causal estimate than *cis*-MR analysis. Although it is possible that the direction of pleiotropy varies at random across the many loci selected and so balances overall (balanced horizontal pleiotropy), this cannot be guaranteed. Although methods are also available to reduce bias due to horizontal pleiotropy in genome-wide MR analysis (e.g., MR-Egger), each relies on other assumptions that may not be fully upheld.[1–4]

### *trans*-weighted *cis*-MR analysis

To illustrate the principle of trans-weighted *cis*-MR analysis of CRP, consider a protein of interest $P_1$ encoded by gene $G_1$ which is *trans*-for-CRP in a GWAS. If $P_1$ is measured directly and it can be used to weight the effect of genetic instruments in $G_1$ that are *cis* for $P_1$ , then the MR estimate of $P_1$ on $D$ is $\frac{(\sigma_{\mathrm{P}_1} \cdot \delta_D^{\mathrm{P}_1}) + \emptyset_D^{\mathrm{GP}_1}}{\sigma_{\mathrm{P}_1}}$, the same as a *cis*-MR. However, if genetic effect estimates for *cis*-variants on $P_1$ are unavailable, then a *cis*-MR analysis of $P_1$ on $D$ can still be undertaken, with the effects of the genetic variants acting in *cis* being weighted by CRP rather than $P_1$. In this case, the MR estimate for the causal effect of $P_1$ on $D$ is *trans*-weighted and given by $\frac{(\sigma_{P1} . \delta_D^{P1}) + (\sigma_{P1} \cdot \delta_{CRP}^{P1} . \delta_D^{CRP}) + \emptyset_D^{G1} + (\emptyset_{CRP}^{\mathrm{G1}} \cdot \delta_D^{CRP})}{(\sigma_{P1} . \delta_{CRP}^{P1})}$ . If $\emptyset_D^{G1} = \emptyset_{CRP}^{\mathrm{G1}} = 0$, the MR estimate for the causal effect of $P_1$ on $D$ is reduced to $\frac{\delta_D^{P1} + \delta_{CRP}^{P1} . \delta_D^{CRP}}{\delta_{CRP}^{P1}}$ . If $\delta_D^{CRP} = 0$ , (i.e., no causal effect of CRP on $D$), then the MR estimate for the causal effect of $P_1$ on $D$ is reduced further to $\frac{\delta_D^{P1}}{\delta_{CRP}^{P1}}$. Thus, in this *trans*-weighted, *cis*-MR analysis, the causal effect of $P_1$ on $D$ is expressed in terms of the causal effect of $P_1$ on CRP. If $\delta_D^{P1}$ and $\delta_{CRP}^{P1}$ are disproportionate, the causal estimate of $P_1$ on $D$ may be inflated or attenuated but still provides a valid test of the null hypothesis, so the association is valid, but its magnitude may not be. The direction of effect in such analyses also requires careful consideration and is discussed in more detail in the next section.

### Empirical validation – consistency between trans- and cis-weighted cis-MR analyses

As noted above, in *trans*-weighted *cis*-MR analysis, the causal effect of a given protein e.g., $P_1$ on $D$ is expressed in terms of the causal effect of $P_1$ on CRP. If $\delta_D^{P1}$ and $\delta_{CRP}^{P1}$ are disproportionate, the causal estimate of $P_1$ on $D$ may be inflated or attenuated but still provides a valid test of the null hypothesis. Where both types of analyses can be undertaken, the naively observed effect direction in a *trans*-weighted *cis*-MR analyses may differ from that in *cis*-weighted *cis*-MR analysis. But the effect directions in the two cases can be resolved as follows. The predicted effect direction of an unobserved *cis*-weighted *cis*-MR estimate ($\delta_D^{P1}$) for $P_1$ on $D$, may be predicted to be equal to the product of the effect directions for (i) the *trans*-weighted estimate, $\frac{\delta_D^{P1}}{\delta_{CRP}^{P1}}$ and (ii) the causal effect of protein $P_1$ on CRP, $\delta_{CRP}^{P1}$ (where sgn $(\bullet)$ is the sign function which extracts the sign of a real number, which is -1 for a negative number, +1 for a positive number, or 0 for the number 0).

$$\mathrm{sgn}\left(\delta_D^{P1}\right) = \mathrm{sgn}\left(\frac{\delta_D^{P1}}{\delta_{CRP}^{P1}}\right) \cdot \mathrm{sgn}(\delta_{CRP}^{P1})$$

We evaluated sgn $\left(\delta_D^{P1}\right)$ (*cis*-weighted *cis*-MR) and sgn$\left(\frac{\delta_D^{P1}}{\delta_{CRP}^{P1}}\right)$ (*trans*-weighted *trans*-MR) for five proteins (275 protein-outcome pairs) and 25 mRNAs (1132 mRNA-outcome pairs) with available protein quantitative trait loci (pQTL) and whole blood expression quantitative trait loci (eQTL) *cis*-MR estimates respectively. We also evaluated *cis*-MR estimates for the causal effects of these proteins and mRNAs on CRP concentration, sgn $(\delta_{CRP}^{P1})$. The predicted and observed effect directions were compared for cases where $\frac{\delta_D^{P1}}{\delta_{CRP}^{P1}}$, $\delta_D^{P1}$ and $\delta_{CRP}^{P1}$ were all significant at varying p value thresholds. At an alpha of 0.05, the observed effect directions for pQTL and whole blood eQTL *cis*-MR analyses matched those predicted for 54 out of 72 (75%) and 172 out of 186 (92%) protein-outcome pairs. The degree of concordance improved with increasingly stringent alpha values (**Mathematical appendix, Figure A1**).

**MENDELIAN RANDOMISATION ANALYSIS OF CRP**

| Exposure of interest | Outcome of interest | Genetic instrument | Exposure used | Causal inference | Type of Mendelian randomisation analysis | Mendelian randomisation estimate | Pleiotropy term | Pleiotropy mitigation | Comment |
|---|---|---|---|---|---|---|---|---|---|
| **CRP** | **D** | $G_{\mathrm{CRP}}$ | CRP | CRP on D | *cis*-MR of CRP (*cis*-weighted) | $\frac{\sigma_{CRP}\delta_D^{CRP} + \emptyset_D^{\mathrm{GCRP}}}{\sigma_{CRP}}$ | $\emptyset_D^{\mathrm{GCRP}}$ | *cis*-MR, colocalization | Preferred approach to estimating the causal effect of a protein (in this example, CRP) on disease because use of genetic instruments in the vicinity of the gene of interest limits opportunities for an effect on disease independent of the encoded protein. Statistical colocalization of variants in the encoding gene that influence protein concentration and those that influence disease risk can reduce the risk of confounding by linkage disequilibrium. If $\emptyset_D^{\mathrm{GCRP}} = 0$, the MR estimate= $\delta_D^{CRP}$, which is the causal effect of CRP on disease. |
| **CRP** | **D** | $G_1$ | CRP | CRP on D | Single locus *trans*-MR of CRP | $\frac{\sigma_{P1}\delta_D^{P1} + \sigma_{P1}\delta_{CRP}^{P1}.\delta_D^{CRP} + \emptyset_D^{G1} + \emptyset_{CRP}^{\mathrm{G1}}\delta_D^{CRP}}{\sigma_{P1}\delta_{CRP}^{P1}}$ | $\sigma_{P1} \cdot \delta_D^{P1} + \emptyset_D^{G1}$ | Limited | High risk of a causal chain independent of the protein of interest, in this example CRP. If $\emptyset_D^{G1} = \emptyset_{CRP}^{\mathrm{G1}} = 0$, the MR estimate= $\frac{\delta_D^{P1}+\delta_{CRP}^{P1}.\delta_D^{CRP}}{\delta_{CRP}^{P1}}$. Only if $\delta_D^{P1} = 0$ (no causal effect of $\mathrm{P}_1$ on D) does the MR estimate equate to $\delta_D^{CRP}$, the causal effect of CRP on D |
| **MENDELIAN RANDOMISATION ANALYSIS OF $\mathbf{P_1}$ 'MR-FISH'** | | | | | | | | | |
| $\mathbf{P_1}$ | **D** | $G_1$ | $\mathrm{P}_1$ | $\mathrm{P}_1$ on D | *cis*-MR of $\mathrm{P}_1$ (*cis*-weighted) | $\frac{\sigma_{\mathrm{P}_1}\delta_D^{\mathrm{P}_1} + \emptyset_D^{\mathrm{GP}_1}}{\sigma_{\mathrm{P}_1}}$ | $\emptyset_D^{\mathrm{GP}_1}$ | *cis*-MR, colocalization | Statistical colocalization of variants in the encoding gene that influence protein concentration and those that influence disease risk can reduce the risk of confounding by linkage disequilibrium. If $\emptyset_D^{\mathrm{GP}_1} = 0$, the MR estimate= $\delta_D^{\mathrm{P}_1}$, which is the causal effect of $\mathrm{P}_1$ on disease. |
| $\mathbf{P_1}$ | **D** | $G_1$ | CRP | $\mathrm{P}_1$ on D | *cis*-MR of $\mathrm{P}_1$ (*trans*-weighted) | $\frac{\sigma_{P1}\delta_D^{P1} + \sigma_{P1}\delta_{CRP}^{P1}.\delta_D^{CRP} + \emptyset_D^{G1} + \emptyset_{CRP}^{\mathrm{G1}}\delta_D^{CRP}}{\sigma_{P1}\delta_{CRP}^{P1}}$ | $\emptyset_D^{G1} + \emptyset_{CRP}^{\mathrm{G1}}\delta_D^{CRP}$ | *cis*-MR, colocalization | Alternative approach to estimating the causal effect of a protein (in this case $\mathrm{P}_1$) on D if the *cis* genetic effect on $\mathrm{P}_1$ has not been estimated directly. Instead, this is proxied by the genetic effect of $\mathrm{G}_1$ on a downstream protein, in this case CRP. This is possible because $\mathrm{G}_1$ is identified as a trans locus in a GWAS of CRP. If $\delta_D^{P1} \neq 0$, $\mathrm{P}_1$ is a common cause of CRP and D and will be confounder of the observed association of CRP and D. If $\emptyset_D^{G1} = \emptyset_{CRP}^{\mathrm{G1}} = 0$, the MR estimate = $\frac{\delta_D^{P1}+\delta_{CRP}^{P1}.\delta_D^{CRP}}{\delta_{CRP}^{P1}}$. If, $\delta_D^{CRP} = 0$ (i.e no causal effect of CRP on D, the MR estimate = $\frac{\delta_D^{P1}}{\delta_{CRP}^{P1}}$. Thus, the causal effect of $\mathrm{P}_1$ on D is expressed in terms of the effect of $\mathrm{P}_1$ on CRP. If $\delta_D^{P1}$ and $\delta_{CRP}^{P1}$ are disproportionate, the effect estimate $\mathrm{P}_1$ on D may be inflated or diminished, but nevertheless provides a valid test of the null hypothesis. |

**Table A1.** Alternative approaches to Mendelian randomisation analysis to evaluate the causal relevance of CRP encoded by $\mathrm{G_{CRP}}$ or a protein $\mathrm{P}_1$ encoded by $\mathrm{G}_1$, a locus identified through a GWAS of CRP – 'MR Fish'.

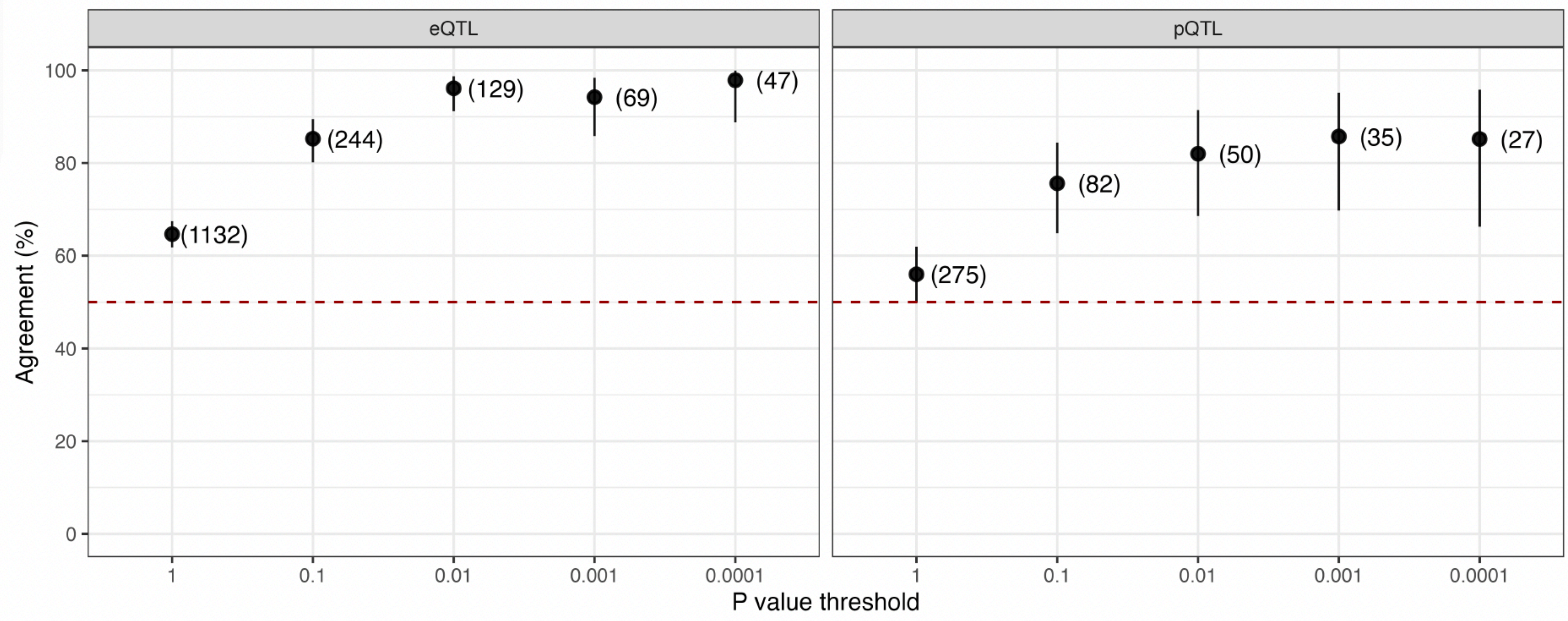


**Figure A1.** Predicted Mendelian randomisation (MR) effect directions for protein-outcome pairs with available CRP-weighted (*trans*-weighted *cis*-MR) and pQTL (*cis*-weighted *cis*-MR) data. For each protein-outcome pair, the causal effect direction (sgn $\left(\delta_D^{P1}\right)$) was predicted to equal the product of the effect directions for (i) *cis*-MR, *trans*-weighted by CRP (sgn$\left(\frac{\delta_D^{P1}}{\delta_{CRP}^{P1}}\right)$), and (ii) the causal effect of the protein on CRP (sgn $(\delta_{CRP}^{P1})$). The degree of concordance (% agreement with 95% confidence intervals) improved when restricting to only protein-outcome pairs with significant MR results for all of sgn $\left(\delta_D^{P1}\right)$, sgn$\left(\frac{\delta_D^{P1}}{\delta_{CRP}^{P1}}\right)$ and sgn $(\delta_{CRP}^{P1})$ at increasingly stringent p values (the total number of protein-outcome pairs meeting each significance threshold is displayed in brackets). All proportions differed significantly from the null hypothesis of 50% (dashed red line) with p values < 0.001. Abbreviations: CRP = C-reactive protein; MR = Mendelian randomisation; eQTL = expression quantitative trait loci; pQTL = protein quantitative trait loci.

**References**


1. Bowden, J., Davey Smith, G. & Burgess, S. Mendelian randomization with invalid instruments: effect estimation and bias detection through Egger regression. *Int. J. Epidemiol.* **44**, 512–525 (2015).
2. Bowden, J., Davey Smith, G., Haycock, P. C. & Burgess, S. Consistent Estimation in Mendelian Randomization with Some Invalid Instruments Using a Weighted Median Estimator. *Genet. Epidemiol.* **40**, 304–314 (2016).
3. Verbanck, M., Chen, C.-Y., Neale, B. & Do, R. Detection of widespread horizontal pleiotropy in causal relationships inferred from Mendelian randomization between complex traits and diseases. *Nat. Genet.* **50**, 693–698 (2018).
4. Foley, C. N., Mason, A. M., Kirk, P. D. W. & Burgess, S. MR-Clust: clustering of genetic variants in Mendelian randomization with similar causal estimates. *Bioinforma. Oxf. Engl.* **37**, 531–541 (2021).
